# Defining diseases from the bottom up with agnostic latent phenotype identification

**DOI:** 10.64898/2026.11.26.26369028

**Authors:** Paul Guerry, Alexandre Fabre

**Affiliations:** Green Grow Scientific, Marseille, France; APHM, Timone Enfants, Pédiatrie Multidisciplinaire, Marseille, France; Aix-Marseille University, INSERM, MMG, Marseille, France

**Keywords:** Human Phenotype Ontology, latent phenotypes, unsupervised clustering, latent class analysis, *MYO5B*, microvillus inclusion disease, large language models

## Abstract

**Background:** Rare diseases are conveniently defined by the causative gene, but whether the corresponding patients form a distinct phenotypic group, or possibly several, is rarely tested.

**Methods:** ALPHI (Agnostic Latent PHenotype Identification) groups patients described by Human Phenotype Ontology terms without reference to diagnostic labels. Distances between patients are embedded with uniform manifold approximation and projection, clustered with a density-based algorithm and checked against a latent class model. ALPHI was evaluated on five sets of simulated cohorts, applied to 137 published patients with *MYO5B*-related disease, and compared with five large language models given the same tasks.

**Results:** In simulated cohorts, ALPHI recognized different combinations of explicit and latent phenotypes (distinct syndromes, distinct diseases, and latent subgroups within or across diseases) with high sensitivity and specificity, and divided only 1 of 1,810 cohorts without structure. In the *MYO5B* cohort, ALPHI identified the three previously suggested phenotypes (microvillus inclusion disease (MVID), progressive familial intrahepatic cholestasis (PFIC) and combined MVID+PFIC), based only on literature-reported symptoms; genotype and survival, which were not used for clustering, differed between them. The five language models failed on at least one and in general most simulated cohorts, and even the best-performing model failed to isolate the MVID+PFIC phenotype. ALPHI is also faster (16–17 s vs ∼1–7 min per cohort) and runs locally, without patient data being sent to an external service.

**Conclusions:** ALPHI provides a reproducible, bottom-up test of whether a proposed disease or subtype exists as a distinct phenotype, and can reveal phenotypes that have not yet been delineated.

## Introduction

Rare diseases were until recently delineated by their phenotype, the set of observable physical and behavioral traits that distinguish these patients from others and from healthy individuals [1]. The phenotype of a disease is a property of a group of patients: phenotypic features are only present in a proportion of patients, making a disease phenotype a statistical object. The Human Phenotype Ontology (HPO) has been widely adopted as a standardized vocabulary to describe phenotypes and to compare them between patients and diseases [2, 3].

The advent of next-generation sequencing has made molecular etiology central to the delineation of Mendelian disorders. A dyadic nomenclature, combining the causative gene with a phenotypic descriptor, has even been proposed [4], although not without dissent [5]. In either case, the phenotypic element rests on expert judgment: whether a phenotype is sufficiently distinct to define a disorder “ultimately reflects the wisdom and judgment of practitioners and researchers in the field” [4]. Such distinctions have rarely been tested statistically. The diagnostic criteria of Alagille syndrome, for instance, were based on the frequency of each abnormality in a series of 80 patients [6], not on their ability to distinguish the syndrome from other causes of cholestasis. The feature frequencies of Orphanet and HPO disease annotations are likewise frequencies in previously reported cohorts. Whether the patients assigned to a disease form a distinct phenotypic group, and whether patients with variants in the same gene form one group or several, remains largely untested.

Two directions can be distinguished in the computational analysis of phenotypes. In top-down analysis, individual patients are compared against the phenotypes of known diseases. Diagnostic tools such as Phenomizer [3] rank known diseases by the semantic similarity of their HPO annotations to the features of a patient. Such tools can only recognize diseases present in their reference annotations, and they inherit the definitions on which these annotations are based. In bottom-up analysis, no disease is defined in advance: patients are compared with one another, and groups emerge from their similarities, from the patient level up to the phenome. Only bottom-up analysis can test whether a proposed disease or subtype exists as a distinct phenotype, or reveal a phenotype that has not yet been delineated.

Previous bottom-up analyses of patients described in HPO codes have involved partitioning a matrix of pairwise semantic similarities with a predefined number of clusters [7, 8]. In these approaches, every patient is assigned to a cluster, and the reliability of individual clusters is seldom assessed, so that the existence of the groups is assumed rather than tested. These analyses were also applied to cohorts annotated by a single team. Cohorts assembled from the published literature, as is often required for rare diseases, are more likely to include features described in different terms by different authors.

It may appear simpler to ask a large language model (LLM) to group published patients. LLMs have been evaluated for phenotype-driven diagnosis of rare diseases [9], but a grouping produced by an LLM is not agnostic. LLMs are trained on the published literature, including the case reports and the classifications of the diseases concerned, and draw on this knowledge whether instructed to or not. Their groupings may therefore reproduce existing classifications rather than test them, even when no disease is named. Results are also prompt-dependent, vary between runs (even with identical prompts), and lack both an explicit distance between patients and any reliability measure.

This article presents ALPHI (Agnostic Latent PHenotype Identification), a software tool to identify the number of distinct phenotypic groups in a cohort of patients based only on sets of HPO terms. ALPHI was evaluated on three sets of simulated cohorts: the 44 distinct syndromes used to evaluate Phenomizer [3], combined in a single cohort; latent subgroups within one disease; and two or three distinct diseases, to estimate the number of patients required. It was then applied to 137 published patients with *MYO5B*-related disease, in which microvillus inclusion disease (MVID), progressive familial intrahepatic cholestasis (PFIC) and combined presentations have been described [10]. The clusters obtained were compared in terms of absent symptoms, genotype and survival, none of which were used for clustering. The same synthetic and *MYO5B* cohorts were also submitted to five LLMs.

## Methods

### Overview of ALPHI

ALPHI (Agnostic Latent PHenotype Identification) clusters the patients of a cohort according to their reported phenotypes, encoded as Human Phenotype Ontology (HPO) terms [2]. A distance is computed between every pair of patients, the distance matrix is embedded with uniform manifold approximation and projection (UMAP [11]), and clusters are identified in the embedding with hierarchical density-based spatial clustering of applications with noise (HDBSCAN [12]) over a grid of parameters. The selected clustering is then checked against a latent class model fitted to the same patients, which replaces it when the density-based clustering has merged groups that the model separates reliably. Each cluster is described by the prevalence of its symptoms and by the symptoms that distinguish it from the rest of the cohort.

### Distances between patients

Three distance measures were compared, each computed on the set of HPO terms reported for each patient. Organ branch (OB) and Resnik distances weight the similarity of terms by their position in the HPO; the Jaccard distance, which ignores the ontology, was used as a baseline. All other steps of ALPHI were identical with the three distances, and the three versions without the latent class check are referred to as ALPHI-OB, ALPHI-Jaccard and ALPHI-Resnik. ALPHI denotes the complete method: the OB distance followed by the latent class check.

### OB distance

The OB distance between two symptoms is the number of steps separating them in the HPO through their closest common ancestor, counted only within the branch of one organ system (a top-level HPO category). Identical terms score −0.5, so that exact matches are favored over near matches, and terms of opposite direction (e.g. increased vs decreased) receive a penalty of 5. Each symptom of one patient is matched to the closest symptom of the other patient within the same organ system, in both directions, and the OB distance is the mean of these best-match distances, floored at 0 so that the distance between two patients is never negative. When an organ system is involved in only one of the two patients, each of its symptoms is matched to the closest symptom of the other patient anywhere in the HPO. Related terms, which arise notably when different authors describe the same feature with different terms, are therefore closer in OB space than unrelated terms.

### Jaccard distance

The Jaccard distance is one minus the intersection size of the two sets (the number of terms shared by two patients) divided by the union size of the two sets (the number of terms reported in either set). The Jaccard index is a standard similarity measure for binary presence/absence data [13] and is used, for example, to compare species occurrences in ecology, as the intersection over union in image segmentation, and as the critical success index in forecast verification.

### Resnik distance

The Resnik distance is one minus the best-match average of Resnik similarities [14], as implemented in Phenomizer [3]. The similarity of two terms is the information content (IC) of their most informative common ancestor. IC values were derived from the HPO disease annotations (phenotype.hpoa, release of 2 September 2026) as −log of the proportion of annotated diseases using the term or one of its descendants, and scaled to 0–1.

All analyses used the same version of the HPO, in which Hepatic fibrosis (HP:0001395) was added as a parent of Portal fibrosis (HP:0006580; this relation has since been incorporated into the official release).

### Clustering and selection

The distance matrix was embedded with UMAP on precomputed distances, with min_dist = 0 and n_neighbors = 30, the settings recommended for clustering. n_neighbors was automatically reduced to 10 for cohorts of 16–50 patients and to N − 2 for cohorts of 15 patients or fewer. The embedding had 10 dimensions for cohorts of 100 patients or more, 3 for 30–99 patients and 2 below 30, with a fixed random seed. HDBSCAN was run with excess-of-mass cluster selection over a grid of minimum cluster sizes (5 to 25 in steps of 5, capped at approximately N/10) and minimum sample numbers (equal to the minimum cluster size or 5 below it). Each configuration was scored by its density-based cluster validity (DBCV [15]) multiplied by the proportion of patients assigned to a cluster, or by this proportion alone when more than half of the configurations had no DBCV. A penalty for local instability, defined as the standard deviation of the number of clusters obtained when both HDBSCAN parameters were varied by ±1, was subtracted. The highest-scoring configuration was retained as the optimal solution. The best configuration for each other number of clusters was reported as an alternative if its score was at least 75% of the optimum.

### Reliability checks

#### Resampling stability

The selected configuration was applied to 50 random subsamples of 80% of the patients [16]. Each cluster was matched to its most similar cluster in each subsample by the Jaccard index of their members. A mean Jaccard index ≥ 0.75 was considered stable, and ≤ 0.5 unsupported.

#### Sparse-feature check

To identify clusters separated by too few symptoms to be considered distinct phenotypes, for each pair of clusters, the symptoms whose prevalence differed by at least 30 percentage points between the two clusters were counted. A cluster pair was flagged sparsely different when no more than two symptoms differed to that extent and the largest difference was at least 50 points. This pairwise design also allowed the detection of a single true group divided by one isolated symptom.

### Latent class check

Density-based clustering separates groups by gaps in the embedding. A subgroup that shares features with two others, such as patients presenting both diseases of a cohort, lies between them and can bridge them into a single cluster. Weakly expressed subgroups can also be absorbed by larger ones. The clustering selected by ALPHI-OB was therefore compared with a latent class model fitted to the same patients. The two methods rest on different principles, distances between patients for ALPHI-OB and a probabilistic model of feature frequencies for the latent class model, so that a structure found by both is unlikely to be an artifact of either.

#### Feature families

The HPO terms reported in the cohort were grouped by complete linkage on their step distance in the HPO (the number of steps through their closest common ancestor), so that any two terms of a family were at most two steps apart (a term, its parent and its grandparent, or siblings). A patient was considered to present a family when reporting any of its terms, so that the same feature described with different terms counted once. Families present in fewer than max(3, 0.03N) patients, or absent in fewer, were not used.

#### Latent class model

A mixture of independent Bernoulli variables was fitted by expectation– maximization, with Beta(1, 1) smoothing of the class-conditional frequencies, ten random starts and one start from the ALPHI clusters (three random starts above ten classes). Models with 1–6 classes and with k − 3 to k + 3 classes, k being the number of ALPHI clusters, were fitted, and the number of classes was chosen by the Bayesian information criterion (BIC). Each patient was assigned to the class of highest posterior probability.

#### Checks of the latent classes

The model with the chosen number of classes was refitted on 20 random subsamples of 80% of the patients, and each class was matched to its most similar class by the Jaccard index of members; classes were considered stable when the mean Jaccard index was at least 0.75. The sparse-feature check was applied to the classes as to the ALPHI clusters.

#### Decision

The ALPHI-OB clusters were kept (i) if the latent classes were as numerous as the ALPHI-OB clusters, whether the two partitions agreed (adjusted Rand index, ARI ≥ 0.8) or not; (ii) if the latent classes were more numerous but at least one was unstable on resampling (mean Jaccard index < 0.75) or flagged by the sparse-feature check; or (iii) if the latent classes were fewer, unless an ALPHI-OB cluster was flagged by the sparse-feature check or unsupported by resampling (mean Jaccard index ≤ 0.5) while the latent classes were stable and unflagged. The latent classes were adopted in the two remaining situations: when they were more numerous than the ALPHI-OB clusters, all stable and none flagged, and when they were fewer and replaced a flagged or unsupported ALPHI-OB cluster.

Patients placed differently by the two methods were listed with their posterior class probabilities. The retained solution was reported as the optimal clustering of ALPHI. In the simulations, the check was evaluated by running ALPHI-OB with and without it on identical cohorts (the same generation seeds).

### Simulation 1: many distinct diseases in one cohort

Simulation 1, the most basic test, assessed the separation of a large number of distinct diseases in a single cohort, following the simulation used to evaluate Phenomizer [3]. This situation arises when patients are retrieved by a phenotype rather than a diagnosis, for example all published patients with congenital limb anomalies, a query that returns patients with Holt–Oram syndrome, split hand–foot malformation and Townes–Brocks syndrome together. Clustering a cohort of many diseases also yields, for each cluster, the symptoms that characterize it and their prevalence, so that phenotype–symptom associations can be derived from patient data, for example to complement curated annotations such as those of the HPO or Orphanet, which are very sparse for some diseases. The 44 syndromes of the Phenomizer study were simulated from the feature frequencies given in its supplementary tables [3]. HPO identifiers were updated to the current release, and 26 identifiers that were absent from it or designated terms unrelated to their label were corrected according to the label. Alternative features (A or B) were represented by their first term, and textual frequencies were converted with the values given by Köhler et al. [3].

Each patient was assigned male or female sex with equal probability, and each feature was drawn independently with its frequency, sex-specific features being restricted to the corresponding sex. Patients drawn without any feature were drawn again (a median of 4 of 880 patients per cohort, range 1–9, mostly with Holt–Oram syndrome). Each cohort comprised 20 patients of each syndrome (880 patients). Noise was added as described by Köhler et al. [3]: for each patient, half as many unrelated HPO terms as features (q = 0.5, rounded down) were drawn at random from the phenotypic abnormality branch, excluding ancestors and descendants of the terms annotated to the syndrome and of each other. Cohorts without noise (q = 0) were also analyzed, and term substitution was not applied. No minimum number of terms per patient was applied, so that all simulated patients were clustered. Ten cohorts were generated for each setting, the same patients being used for each distance and noise level. A syndrome was considered recovered when one cluster contained at least half of its patients and the syndrome formed a majority of that cluster.

### Simulation 2: latent subgroups within one disease

Simulation 2 addressed the more challenging task of identifying latent subgroups within a single diagnosis: patients in the cohort share a set of core symptoms and differ by only a few features. This is the situation of a disease whose phenotypic spectrum is incompletely described, for example *MYO5B* deficiency, first described as microvillus inclusion disease and later found also to cause cholestasis, sometimes without intestinal disease. Each simulated cohort comprised patients with one artificial disease defined by 20 core symptoms, each present at a frequency taken from the Orphanet listing of the disease. Latent subgroups were defined by signature sets of Δs distinguishing symptoms, each present in 80% of the patients of the subgroup itself, and in 5% of patients in the rest of the cohort. Patients were divided evenly between subgroups. Three scenarios were simulated:

- **D1S0**: one disease (D1) without subgroups (S0). The signature symptoms of three sets were present at a uniform 30%, giving the same overall frequencies as in D1S3, but without any structure: any split in these cohorts is therefore a false positive. This situation represents a disease with variable expressivity but no subgroup structure.
- **D1S2**: one disease (D1) with two latent subgroups (S2).
- **D1S3**: one disease (D1) with three latent subgroups (S3).

The simulation grid combined cohort size (N = 30, 90, 180, 360), number of distinguishing symptoms (Δs = 3, 6, 9), distribution of the distinguishing symptoms (dispersed across organ systems or concentrated in one), and term substitution, with substitution probability *p* (0, 0.25, 0.5). Substitution was modeled by replacing each reported term by its parent (two-thirds of cases) or a sibling term (one third), mimicking the heterogeneous vocabulary of case reports. The grid was repeated for three compositions (C1–C3), each of 65 symptoms (20 core and 45 signature symptoms), drawn from the pooled Orphanet symptom lists of 14 diseases (Table S1) with no symptom shared between compositions, to avoid dependence on a single vocabulary. Five cohorts were generated for each setting (3,240 cohorts).

Each variable of the grid (Δs, *p* and *r*) represents a different source of noise in published cohorts. The number of distinguishing symptoms, Δs, accounts for variations in the number of features by which subgroups differ. The level of term substitution, *p*, accounts for variations in the terms used by different authors to describe the same symptom. The level of reporting, *r*, accounts for variations in the number of present features actually reported for each patient.

Each grid point was simulated 5 times, and 10 times when the result lay within two standard errors of a threshold. Generation seeds depended only on the cohort, so that comparisons between layouts, substitution levels and distances were made on identical patients. The primary outcome was recovery of the true number of subgroups by the optimal solution; agreement with the true subgroups was measured by the ARI.

Incomplete reporting was investigated in an additional set of cohorts (composition C1, dispersed layout, Δs = 6, *p* = 0.25, N = 90 and 180, all three scenarios), in which each distinguishing symptom was reported with probability *r* = 0.5.

### Simulation 3: two or three distinct diseases in one cohort

Simulation 3 assessed the number of patients required to separate two or three diseases known to be distinct. This situation may occur in practice when one gene causes clinically distinct disorders, for example LMNA variants, which cause muscular dystrophy, lipodystrophy and progeria, or when a cohort assembled around a shared feature contains patients with several diseases. Simulated patients with Marfan syndrome, Wilson disease and Alagille syndrome were generated using Orphanet symptom lists. For every cohort, the frequency of each symptom was drawn uniformly within its Orphanet frequency class (e.g. frequent, 30–79%). Patients were assigned male or female sex in equal proportions, and sex-specific symptoms were restricted to the corresponding sex.

Two-disease cohorts were generated with in a 2:1 ratio of the corresponding patients (e.g. two Wilson patients for each Alagille patient, and vice versa), and three-disease cohorts likewise, with a 3:2:1 ratio of patient numbers (e.g. three Marfan patients for every two Alagille patients and one Wilson patient), in three rotations. Single-disease cohorts (N = 10, 30, 90, 180) were also generated as negative controls. Reporting (each symptom reported with probability *r* = 1 or 0.5, since published patient descriptions list far fewer symptoms than are listed in the Orphanet summaries) and term substitution (*p* = 0, 0.25, 0.5) were also varied. For each scenario and setting, the smallest cohort size at which the true number of diseases was recovered in at least 80% of runs was determined by bisection on a ladder of 10, 20, 30, 45, 60, 90, 120 and 180 patients.

### Simulation 4: a combined subgroup and a marker with two causes

Simulation 4 reproduced two features of phenotypic structure that density-based clustering handles poorly: a subgroup presenting the features of two others, and a feature with two causes. The combined subgroup represents patients presenting both of the disorders caused by one gene, for example *MYO5B* patients with both microvillus inclusion disease and cholestatic liver disease. The marker with two causes represents a feature that can belong to one subgroup’s disease but arise in another subgroup through a different mechanism, for example cholestasis caused by parenteral nutrition in patients with microvillus inclusion disease, rather than by the *MYO5B* liver disease itself. Each cohort comprised 140 patients in three subgroups of 70 (A), 45 (B) and 25 (AB), assigned in random order, with the 20 core symptoms of composition C1. Subgroups A and B were defined by signature sets of six symptoms, each present in 80% of patients of the subgroup and 5% of the others; subgroup AB presented both signature sets and, in half of the settings, three further symptoms of its own (present in 80% of AB patients and 5% of the others). One symptom of set B was also present in 30% of the patients of A (*f* = 0.3) without the rest of set B, as a feature acquired through another mechanism; these patients belonged to A by construction. The grid combined the marker frequency (*f* = 0 or 0.3), the number of AB-specific symptoms (0 or 3) and reporting level (*r* = 1 or 0.75), with term substitution *p* = 0.1, and five cohorts were generated per setting. All methods were compared on identical patients.

The outcomes considered were recovery of the three subgroups, the ARI with the true subgroups, whether AB formed a cluster of its own, and the proportion of marker carriers of A placed in the main cluster of A.

### Simulation 5: latent subgroups crossing known syndromes

Simulation 5 represented the detection of phenotypes that do not correspond to a known diagnosis: two latent subgroups shared across two well-characterized syndromes. This represents a subphenotype shared by patients with different diagnoses and therefore invisible to a grouping by diagnosis, for example a complication of a common treatment, or the effect of a modifier gene or an environmental exposure present in patients with several syndromes. Each cohort comprised two latent subgroups of 75 (G1) and 50 (G2) patients, defined by distinguishing sets of six symptoms with no common clinical interpretation (G1: migraine, hypothyroidism, acne, varicose veins, tinnitus and recurrent urinary tract infections; G2: atopic dermatitis, insomnia, hyperuricemia, vitiligo, hearing impairment and gastroesophageal reflux). Each distinguishing symptom was absent outside its subgroup and present within it with a frequency drawn around 80% (normal, SD 0.08, bounded by 0.55 and 0.95). Independently of the latent subgroup, each patient was given the appearance of Marfan syndrome or Noonan syndrome with equal probability, through four features of that syndrome, each present with probability 0.7 (Marfan: ectopia lentis, arachnodactyly, aortic root aneurysm, dural ectasia; Noonan: webbed neck, pulmonic stenosis, hypertrophic cardiomyopathy, ptosis). The true structure therefore comprised four groups: two latent subgroups within both syndromes.

Reporting was made to resemble published case series. Patients were assigned to 12 series of unequal size (Dirichlet weights), each with its own typical number of reported terms; each patient’s number of terms was drawn around that of the series and bounded by 8 and 20. Patients with fewer terms were completed with common findings unrelated to either structure (21 terms such as fever, cough, recurrent otitis media and constipation, weighted by commonness); signature and syndrome features were never removed. Five cohorts were generated. Outcomes were the ARI with the four true groups and, separately, with the latent subgroups alone and with the syndromes alone, so that a solution reproducing only one of the two factors could be identified.

### Comparison with language models

Five LLMs were given the same grouping tasks through their public chat interfaces in November 2026: Claude (Opus 5.5, highest effort setting), ChatGPT (reported by the model as GPT-6, Think mode), Gemini (Auto mode, Gemini 3 models, high thinking level), Grok (Grok 4.7, Expert mode) and DeepSeek (version not displayed, deep thinking mode). Each model was thus used at its highest available reasoning setting. Each prompt was submitted in a new private or temporary chat (for DeepSeek, which has no such mode, in a new chat), with web search disabled where the interface allowed it. The prompt asked for the patients to be grouped according to their phenotype, based only on their reported symptoms; it stated that the number of groups was not known in advance and might be one, excluded the use of the web or of any external source, and requested one row per patient in a comma-separated values (CSV) table. Patients were identified by random codes, presented in random order, and described by the labels of their reported HPO terms, listed in random order. Prompts were pasted into the chat window, except for Gemini, whose input field truncates text beyond approximately 32,000 characters; for Gemini, the prompt was uploaded as a text file with the message “Follow the instructions in the attached file.” All prompts have been uploaded to the ALPHI repository (simulations/llm_prompts).

Four settings of Simulation 2 were used, with dispersed signature symptoms and composition C1: D1S2 with N = 90, Δs = 6 and *p* = 0.25; D1S2 with N = 180, Δs = 3 and *p* = 0; D1S3 with N = 180, Δs = 6 and *p* = 0.25; and D1S0 with N = 180, Δs = 6 and *p* = 0.25. The five cohorts of each setting were regenerated from the seeds recorded in the simulation grid, so that the models were given the same patients as ALPHI, and each cohort was submitted once to each model. A cohort was considered resolved when the number of groups returned equaled the true number of subgroups and the ARI with the true subgroups (for D1S2 and D1S3 only) was at least 0.8; the same criterion was applied to the optimal ALPHI solutions.

The five cohorts of Simulation 5 were submitted once to each model in the same way, without any gene or disease named. Agreement was measured by the ARI with the four true groups, with the latent subgroups and with the syndromes, as for ALPHI on the same cohorts.

For every answer, the time from submission to the end of the answer was recorded, and patients missing from the returned table were counted. Agreement was computed on the patients returned and, as for patients left unassigned by ALPHI, with missing patients counted as a group of their own.

The 137 patients of the *MYO5B* cohort (see below) were submitted five times to each model under three conditions: without any gene or disease named (blind); with the statement that all patients carried pathogenic variants in *MYO5B* (gene); and with, in addition, the DOI and year of the article reporting each patient (article). Patient order was reshuffled between runs. In the absence of a ground truth, groupings were compared with the ALPHI-OB clusters (ARI) and between runs of the same model and condition (mean pairwise ARI). A simulated D1S0 cohort of 137 patients (Δs = 6, *p* = 0.1), presented in the same way, was submitted five times to each model as a negative control. Answers were screened for mentions of external sources (URLs, DOIs, author names, web searches) and, in the blind and control conditions, for disease labels that the prompt did not give.

### Application to *MYO5B*-related disease

Patients were drawn from PytheasDB [17, 18], an open-access database of published clinical data on rare pediatric digestive diseases, which contained 1,784 patients from 375 articles, with phenotypes coded in HPO. Of the 210 patients with *MYO5B* listed as their gene, including 53 diagnosed clinically without genetic testing, the 137 with at least eight reported HPO terms were analyzed. They originated from 35 articles published between 1978 and 2026, with a median of 11 terms per patient (range 8–24) and 187 distinct terms.

Clusters were interpreted solely through their phenotypic composition; the disease labels recorded in the database were not used. Two signatures were defined from the expected presentations, each term being counted with its descendants: the MVID signature (diarrhea, microvillus inclusions and dependency on parenteral nutrition) and the PFIC signature (cholestasis, pruritus, hyperbilirubinemia, and elevated aspartate or alanine aminotransferase).

Symptoms explicitly reported as absent, which were not used for clustering, were compared between clusters as an independent check of their phenotypic composition. A symptom was considered absent in a patient when it was listed as absent and not as present; intermittent symptoms, recorded as both present and absent in PytheasDB, were excluded. Genotypes and survival, which were not used for clustering either, were also compared between clusters. Variants were classified as null (nonsense, frameshift or splice site) or missense, and genotypes as biallelic null, null with missense, or biallelic missense. The distribution of genotype classes was compared with a permutation test. Survival from birth was estimated by the Kaplan–Meier method and compared with the log-rank test.

The heterogeneity of term use in the *MYO5B* cohort was estimated as a counterpart of the substitution probability *p* of Simulation 2. Within each cluster, terms were taken in decreasing order of frequency, and each term not yet assigned defined a feature family together with the unassigned terms one step away from it in the HPO (parents, children and siblings). A feature was considered coded with a substitute when a patient reported a term of the family but not its most frequent term. The estimator was calibrated on simulated D1S2 cohorts of 136 patients (*s* = 6; five cohorts at each of *p* = 0, 0.1, 0.25, 0.4 and 0.5). The relationship between age at last report and phenotype was examined by comparing cluster membership between age groups (< 1, 1–5 and > 5 years), and by correlating age with the mean ALPHI-OB distance of each patient to the patients of each other cluster (Spearman correlation).

### Software and code availability

ALPHI is written in Python 3.14 and uses umap-learn (v. 0.5.12) and hdbscan (v. 0.8.44). Figures were produced in R 4.5.2 with ggplot2. ALPHI is open-source software released under the MIT licence and is available on GitHub [19] and archived at Zenodo [20], together with the simulation scripts, the HPO-derived files and the PytheasDB export analyzed in this study. Simulation results and LLM prompts are archived at Zenodo [21]. A browser version is available [22]; all computations are performed in the user’s browser, and no patient data are transmitted. Run times were measured on a Dell Tower ECT1250 (Intel Core i7-14700, 20 cores, 28 logical processors) under Windows Subsystem for Linux 2 with 7.6 GB of memory allocated, using 8 worker processes for the computation of distances.

## Results

### Simulation 1: many distinct diseases

The 44 syndromes were separated in a single cohort by all three versions of ALPHI (Table 1). Simulated patients had a mean of 10.8 features (range 1–35), and at least 98% of patients were assigned to a cluster in every cohort. ALPHI-OB and ALPHI-Jaccard recovered a mean of 43.3 of the 44 syndromes without noise, and 42.8 and 43.2 with noise. The exact number of 44 clusters was found in 5 or 6 of 10 cohorts; in the other cohorts, the 44-cluster solution was reported as an alternative in 16 of 17 cases. ALPHI-Resnik recovered fewer syndromes (42.1 without noise, 39.9 with noise) and reported the 44-cluster solution as an alternative in 2 of its 15 incorrect cohorts.

**Table 1.** Separation of the 44 syndromes of Simulation 1

| Noise (q) | Version | Cohorts with 44 clusters | Clusters (range) | Syndromes recovered, mean (min) | Mean ARI |
|---|---|---|---|---|---|
| 0 | ALPHI-OB | 6 | 43–44 | 43.3 (42) | 0.981 |
| 0 | ALPHI-Jaccard | 6 | 43–44 | 43.3 (42) | **0.986** |
| 0 | ALPHI-Resnik | 3 | 41–44 | 42.1 (39) | 0.963 |
| 0.5 | ALPHI-OB | 5 | 41–44 | 42.8 (41) | 0.970 |
| 0.5 | ALPHI-Jaccard | 6 | 42–44 | 43.2 (41) | **0.985** |
| 0.5 | ALPHI-Resnik | 2 | 37–44 | 39.9 (34) | 0.902 |

Results are reported for 10 cohorts of 880 patients per setting, as the number of cohorts with 44 clusters, the range of the number of clusters, the mean (minimum) number of syndromes recovered and the mean ARI with the syndromes. Abbreviations: ARI, adjusted Rand index; q, noise level (unrelated terms added per patient, as a proportion of its features).

Errors consisted almost exclusively of merges of two syndromes into one cluster; a syndrome was divided between clusters in 3 of 2,640 syndrome-cohort combinations, all with ALPHI-Resnik and noise. The merged syndromes shared clinical features: neurofibromatosis types 1 and 2; ectrodactyly, ectodermal dysplasia and cleft lip/palate syndrome 1 (EEC1) and split-hand/foot malformation with long bone deficiency 1 (SHFLD1), both with split hand/foot; distal arthrogryposis type 1 and congenital contractural arachnodactyly, joined by Freeman–Sheldon syndrome with ALPHI-Resnik; and Holt–Oram syndrome and kyphomelic dysplasia. Twenty-seven syndromes were recovered in all 60 cohorts. The syndromes recovered least often were those with the fewest features per patient: SHFLD1 (2.8 features per patient, recovered in 70% of cohorts) and Holt–Oram syndrome (2.0 features, 77%; Table S2).

Noise had little effect on ALPHI-Jaccard (mean ARI 0.986 without and 0.985 with noise) or ALPHI-OB (0.981 and 0.970), but reduced the performance of ALPHI-Resnik (0.963 and 0.902). In paired comparisons, ALPHI-Jaccard obtained a higher ARI than ALPHI-OB in 7 of 10 cohorts without noise and 8 of 10 with noise, and ALPHI-OB a higher ARI than ALPHI-Resnik in 7 and 8 of 10.

### Simulation 2: latent subgroups

With ALPHI, the true number of subgroups was recovered in 96% of the cohorts with two latent subgroups and 87% of those with three subgroups, and none of the 1,080 cohorts without subgroups was divided (Figure 1). With Δs = 6/26 or 9/29, two subgroups were recovered in every cohort and three in every cohort of 90 patients or more; with Δs = 3/23, about 90 patients were needed for two subgroups and 180 for three. The three distances, compared without the latent class check, are described in the rest of this section (Figures S1 and S2).

**Figure 1.**
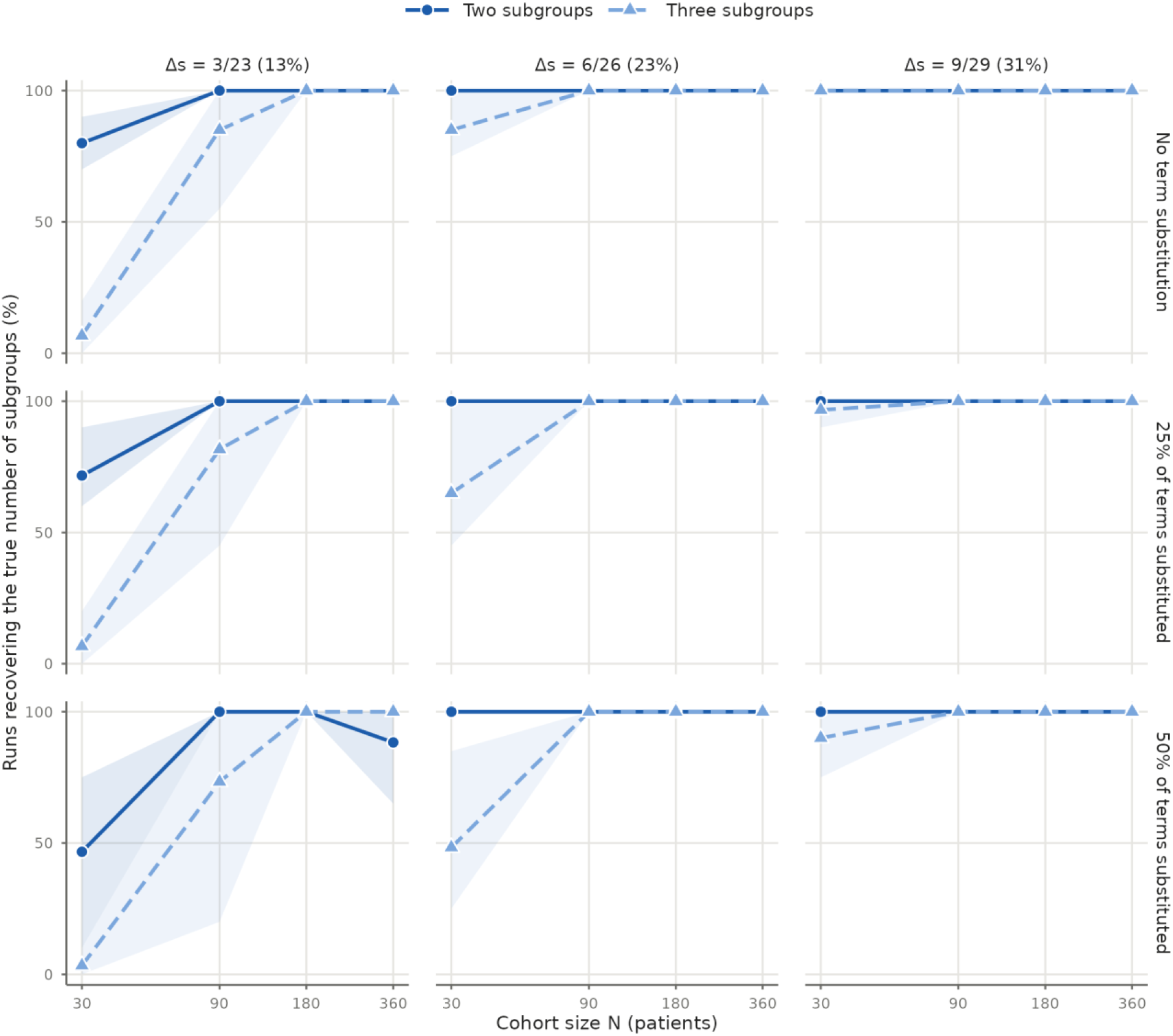
Recovery of latent subgroups by ALPHI (Simulation 2). Percentage of cohorts in which the true number of subgroups was found, by cohort size N, distinguishing symptoms (Δs, columns) and term substitution (rows), for two (solid line) and three (dashed line) latent subgroups. Δs: number of distinguishing symptoms of a subgroup over the number of symptoms defining it (20 core symptoms shared by all subgroups plus the distinguishing symptoms), with the percentage in parentheses. Lines: mean over compositions C1–C3; shaded bands: range over compositions. Dispersed and concentrated layouts are pooled; complete reporting. None of the 1,080 cohorts without subgroups was divided.

Spurious subgroups were identified in just 1 of 1,080 D1S0 cohorts with ALPHI-OB (0.09% of cases), and none of 1,080 with ALPHI-Jaccard or ALPHI-Resnik. In cohorts with subgroups, errors were almost exclusively merges, with fewer clusters than true subgroups.

The most effective version of ALPHI depended on the consistency of the terms used (Table 2; Figures S1 and S2). With exact terms (p = 0), subgroups were recovered most often by ALPHI-Jaccard, which outperformed ALPHI-OB in 91 paired comparisons and was outperformed in six (ARI difference > 0.05). In the presence of any level of term substitution, the performance of ALPHI-Jaccard declined sharply (since for the Jaccard distance, related terms receive no credit). ALPHI-OB was then the most reliable version, outperforming ALPHI-Jaccard in 565 paired comparisons (1 in the reverse direction) and outperforming ALPHI-Resnik in 222 paired comparisons (36 in the reverse direction).

**Table 2.**
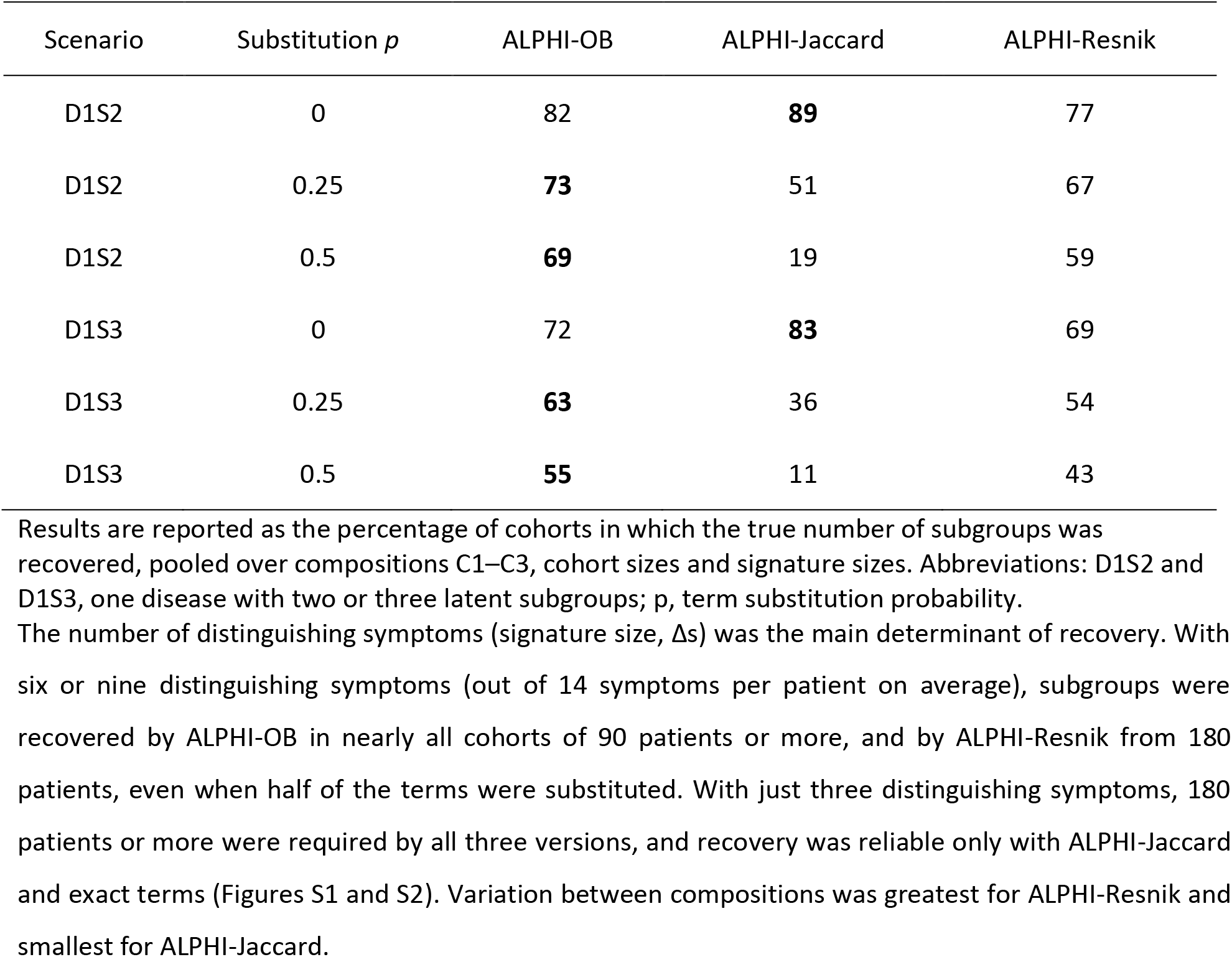
Recovery of the true number of subgroups in Simulation 2

| Scenario | Substitution *p* | ALPHI-OB | ALPHI-Jaccard | ALPHI-Resnik |
|---|---|---|---|---|
| D1S2 | 0 | 82 | **89** | 77 |
| D1S2 | 0.25 | **73** | 51 | 67 |
| D1S2 | 0.5 | **69** | 19 | 59 |
| D1S3 | 0 | 72 | **83** | 69 |
| D1S3 | 0.25 | **63** | 36 | 54 |
| D1S3 | 0.5 | **55** | 11 | 43 |

Results are reported as the percentage of cohorts in which the true number of subgroups was recovered, pooled over compositions C1–C3, cohort sizes and signature sizes. Abbreviations: D1S2 and D1S3, one disease with two or three latent subgroups; p, term substitution probability.

The number of distinguishing symptoms (signature size, Δs) was the main determinant of recovery. With six or nine distinguishing symptoms (out of 14 symptoms per patient on average), subgroups were recovered by ALPHI-OB in nearly all cohorts of 90 patients or more, and by ALPHI-Resnik from 180 patients, even when half of the terms were substituted. With just three distinguishing symptoms, 180 patients or more were required by all three versions, and recovery was reliable only with ALPHI-Jaccard and exact terms (Figures S1 and S2). Variation between compositions was greatest for ALPHI-Resnik and smallest for ALPHI-Jaccard.

Over-splitting occurred in four cohorts with ALPHI-OB, all in composition C2 and in the same setting (D1S2, N = 360, Δs = 3, dispersed layout; two with exact terms and two with p = 0.25), and in none with ALPHI-Jaccard or ALPHI-Resnik. In each of the six such cohorts regenerated at *p* = 0, one true subgroup was divided by a single symptom, thin glomerular basement membrane, present in nearly all patients of one part and absent from the other. This core symptom is frequent but often the only symptom of its organ system in a given patient. All six over-splits were flagged by the sparse-feature check, whereas no pair of true subgroups, which differed on six symptoms, was flagged.

### Simulation 3: two or three distinct diseases

With ALPHI, every pair of diseases (2:1) was separated from 10 patients with complete reporting and from 10–20 patients with half of the symptoms reported, and the three diseases (3:2:1) from 20–45 and 45–120 patients, respectively (Figure 2). One of 370 single-disease controls was divided with complete reporting, and none of 360 with half of the symptoms reported. The three distances, compared without the latent class check, are described in the supplementary information (Figure S3).

**Figure 2.**
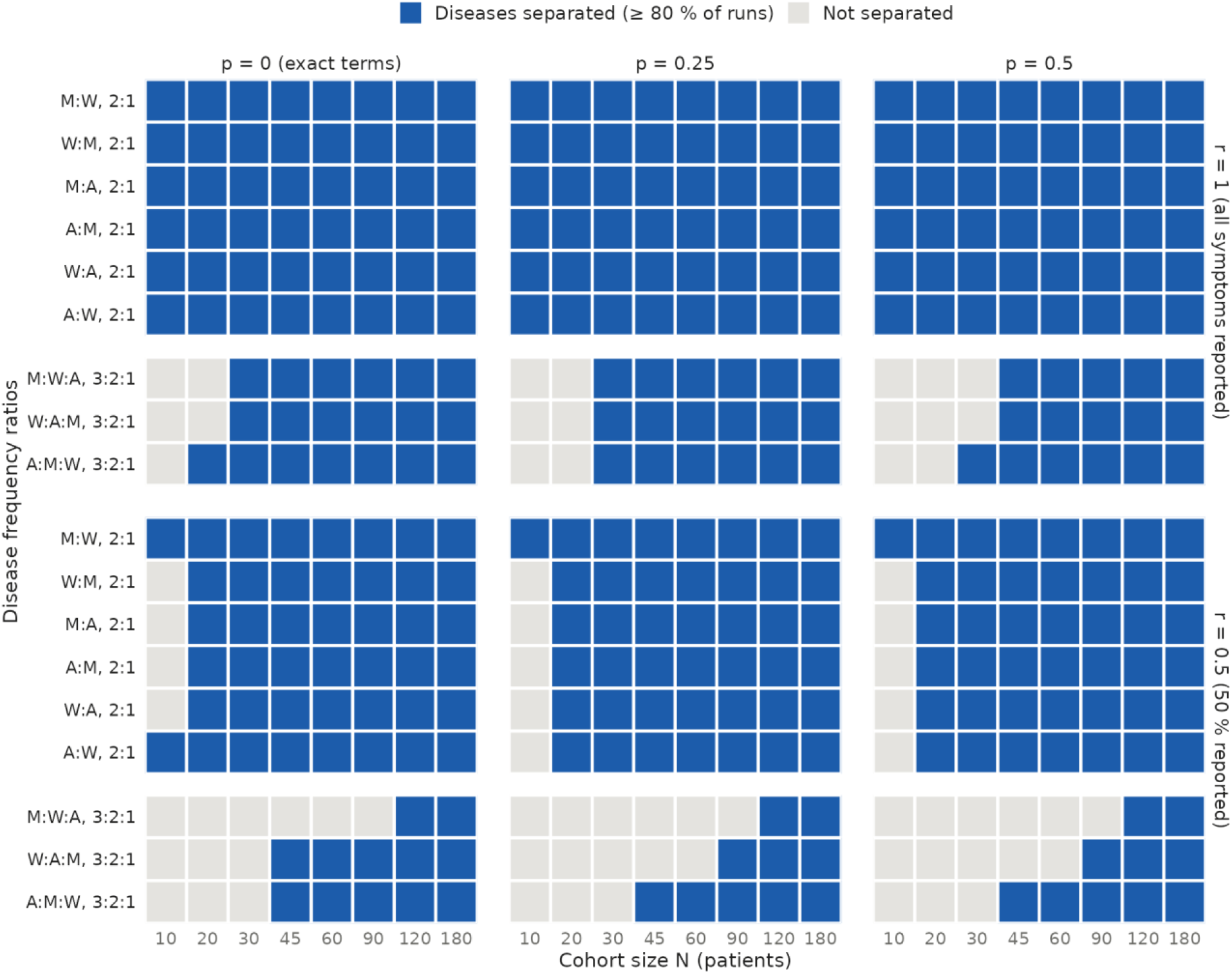
Number of patients required for ALPHI to separate distinct diseases (Simulation 3). For each pair (2:1) or triple (3:2:1) of Marfan syndrome (M), Wilson disease (W) and Alagille syndrome (A), blue tiles mark the cohort sizes at or above the smallest size at which the true number of diseases was found in at least 80% of runs, and grey tiles the sizes below; by reporting rate (*r*, rows) and term substitution (*p*, columns). The smallest size was found by bisection among the sizes shown; separation was assumed to hold at larger sizes once reached. Single-disease controls were divided in 1 of 370 runs with complete reporting and in none of 360 with half of the symptoms reported.

No cohort combining two or three diseases was divided into more clusters than diseases, with any of the three versions of ALPHI (approximately 5,900 cohorts). The number of patients required was determined mainly by the size of the smallest group (Figure S3): no version separated a 2:1 pair with fewer than 20 patients, corresponding to approximately seven patients in the smaller group, or the triplet with fewer than 45 patients.

With complete reporting (r = 1), most scenarios involved a sufficient number of patients for all three versions to separate the subgroups (Table 3); ALPHI-Resnik was the best or equal-best in all 27 settings, ALPHI-OB in 25 and ALPHI-Jaccard in 22. With half of the symptoms reported (r = 0.5), the most sensitive version was ALPHI-Resnik (best or equal-best in 25 of 27 settings), followed by ALPHI-OB (21) and ALPHI-Jaccard (11). ALPHI-Jaccard was again the most affected by substitution.

**Table 3.** Patients required to separate the diseases in Simulation 3

| Reporting *r* | Substitution *p* | ALPHI-OB | ALPHI-Jaccard | ALPHI-Resnik |
|---|---|---|---|---|
| 1 | 0 | 28 | 28 | 26 |
| 1 | 0.25 | 28 | 31 | 26 |
| 1 | 0.5 | 26 | 32 | 26 |
| 0.5 | 0 | 33 | 36 | 30 |
| 0.5 | 0.25 | 36 | 51 | 35 |
| 0.5 | 0.5 | 41 | 78 | 35 |

Results are reported as the geometric mean, over the nine scenarios, of the number of patients required to separate the diseases. Abbreviations: p, term substitution probability; r, reporting probability.

ALPHI-OB performed least well in scenarios combining Wilson disease and Alagille syndrome, two liver diseases with many terms in common. With half of the symptoms reported, 30 to 90 patients were required by ALPHI-OB to separate Wilson:Alagille 2:1 cohorts compared with just 20 by ALPHI-Resnik, and 180 patients were required for Marfan:Wilson:Alagille 3:2:1 triplets, compared with 90 to 120 by ALPHI-Resnik.

In single-disease controls, no cohort was split by ALPHI-Jaccard or ALPHI-Resnik (0 of 720 each). With ALPHI-OB, 6 of 730 cohorts were split, all Wilson disease cohorts of 180 patients with complete reporting and exact terms. Each split separated patients according to the presence of a single symptom that is often the only sign of its organ system: splenomegaly, aminoaciduria or susceptibility to bruising. Five of the six splits were flagged by the sparse-feature check; the sixth was based on three symptoms (aminoaciduria, kidney stones and a menstrual symptom), exceeding the two-symptom limit of the check.

### Effect of the latent class check

The latent class check increased the recovery of true subgroups without introducing spurious ones. In Simulation 2, ALPHI-OB was run with and without the check on the same 3,240 cohorts (compositions C1–C3; Table 4; Figures S4 and S5).

**Table 4.**
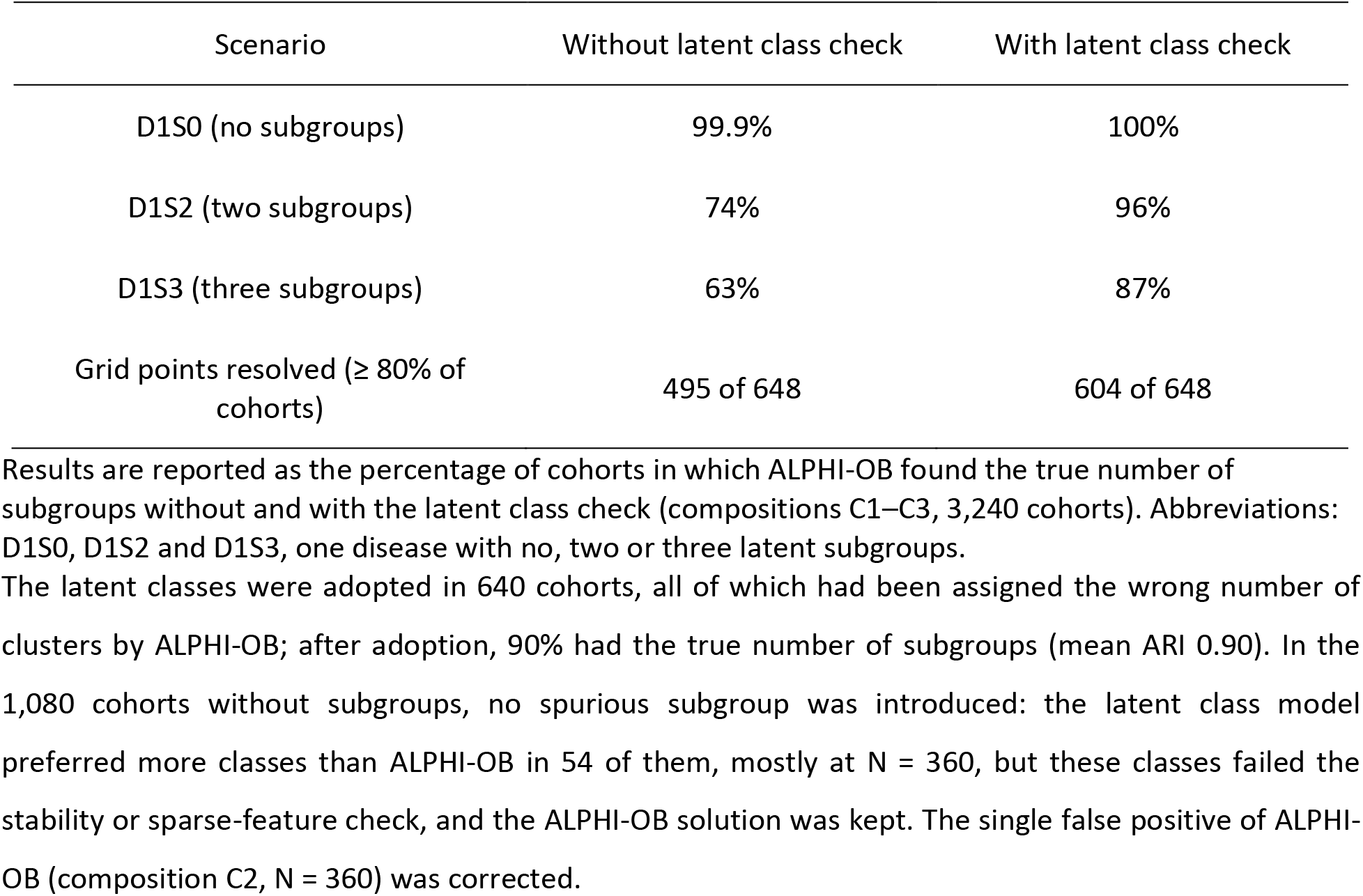
Effect of the latent class check in Simulation 2

| Scenario | Without latent class check | With latent class check |
|---|---|---|
| D1S0 (no subgroups) | 99.9% | 100% |
| D1S2 (two subgroups) | 74% | 96% |
| D1S3 (three subgroups) | 63% | 87% |
| Grid points resolved (≥ 80% of cohorts) | 495 of 648 | 604 of 648 |

Results are reported as the percentage of cohorts in which ALPHI-OB found the true number of subgroups without and with the latent class check (compositions C1–C3, 3,240 cohorts). Abbreviations: D1S0, D1S2 and D1S3, one disease with no, two or three latent subgroups.

The latent classes were adopted in 640 cohorts, all of which had been assigned the wrong number of clusters by ALPHI-OB; after adoption, 90% had the true number of subgroups (mean ARI 0.90). In the 1,080 cohorts without subgroups, no spurious subgroup was introduced: the latent class model preferred more classes than ALPHI-OB in 54 of them, mostly at N = 360, but these classes failed the stability or sparse-feature check, and the ALPHI-OB solution was kept. The single false positive of ALPHI-OB (composition C2, N = 360) was corrected.

The largest gains were obtained where density-based clustering merged subgroups: with three signature symptoms, the true number of subgroups was recovered in 89% of D1S2 cohorts (27% without the check) and 72% of D1S3 cohorts (14%), and with substitution *p* = 0.5, in 94% of D1S2 cohorts (69%). The remaining failures were almost all cohorts of 30 patients with signatures of three or six symptoms, that is, subgroups of 10–15 patients.

With incomplete reporting (*r* = 0.5), ALPHI-OB did not detect any subgroups. With the check, both subgroups of D1S2 were recovered in all 10 cohorts (ARI 0.81 at N = 90 and 0.85 at N = 180), and three subgroups in 5 of 10 D1S3 cohorts of 180 patients; none was found in D1S3 cohorts of 90 patients, and D1S0 cohorts remained undivided.

In Simulation 3, two diseases were separated from 10 patients with the check, compared with 20 without it, and three diseases from 20–45 patients, compared with 45–90. Spurious splits of single-disease cohorts decreased from 6 to 1 of 370 (Figure S6).

In Simulation 1, the check did not modify the ALPHI-OB solution in any of the 20 cohorts, with or without noise terms (Table 1 therefore applies to ALPHI as well). With 44 groups of 20 patients, the latent class model selected fewer classes than there were syndromes, and the ALPHI-OB clusters were kept by the decision rules. The check is therefore neutral for cohorts made of many small groups, and the merges of similar syndromes by ALPHI-OB (for example neurofibromatosis types 1 and 2) remained.

Latent class analysis alone would not have replaced ALPHI-OB. In the 1,080 Simulation 2 cohorts without subgroups, it divided 54 (5%), 49 of them among the 270 cohorts of 360 patients (18%), whereas the combined method divided none, extra classes being adopted only when they passed the stability and sparse-feature checks. In cohorts with subgroups, it recovered the true number of subgroups about as often as the combined method (95% of D1S2 and 82% of D1S3 cohorts), but in no cohort of Simulation 2 was it right where the combined method was wrong, whereas the reverse occurred in 84 cohorts. In Simulation 1, latent class analysis alone failed: when all numbers of classes from 1 to 50 were fitted, the BIC was lowest at 6–10 classes for the 44 syndromes (ARI with the syndromes 0.09–0.16; Table S3). In Simulation 3, latent class analysis alone and the combined method gave the same numbers of groups.

### Combined subgroups and a marker with two causes

A subgroup presenting the features of two others was rarely recovered by density-based clustering, and the latent class check recovered it in most cohorts with complete reporting. Without the check, ALPHI-OB found the three subgroups in 4 of the 40 cohorts of Simulation 4 and merged them into a single cluster in 29; the combined subgroup AB bridged A and B. With the check, the three subgroups were recovered in 17 of 20 cohorts with complete reporting, including 7 of 10 in which AB had no symptom of its own (Table 5).

**Table 5.**
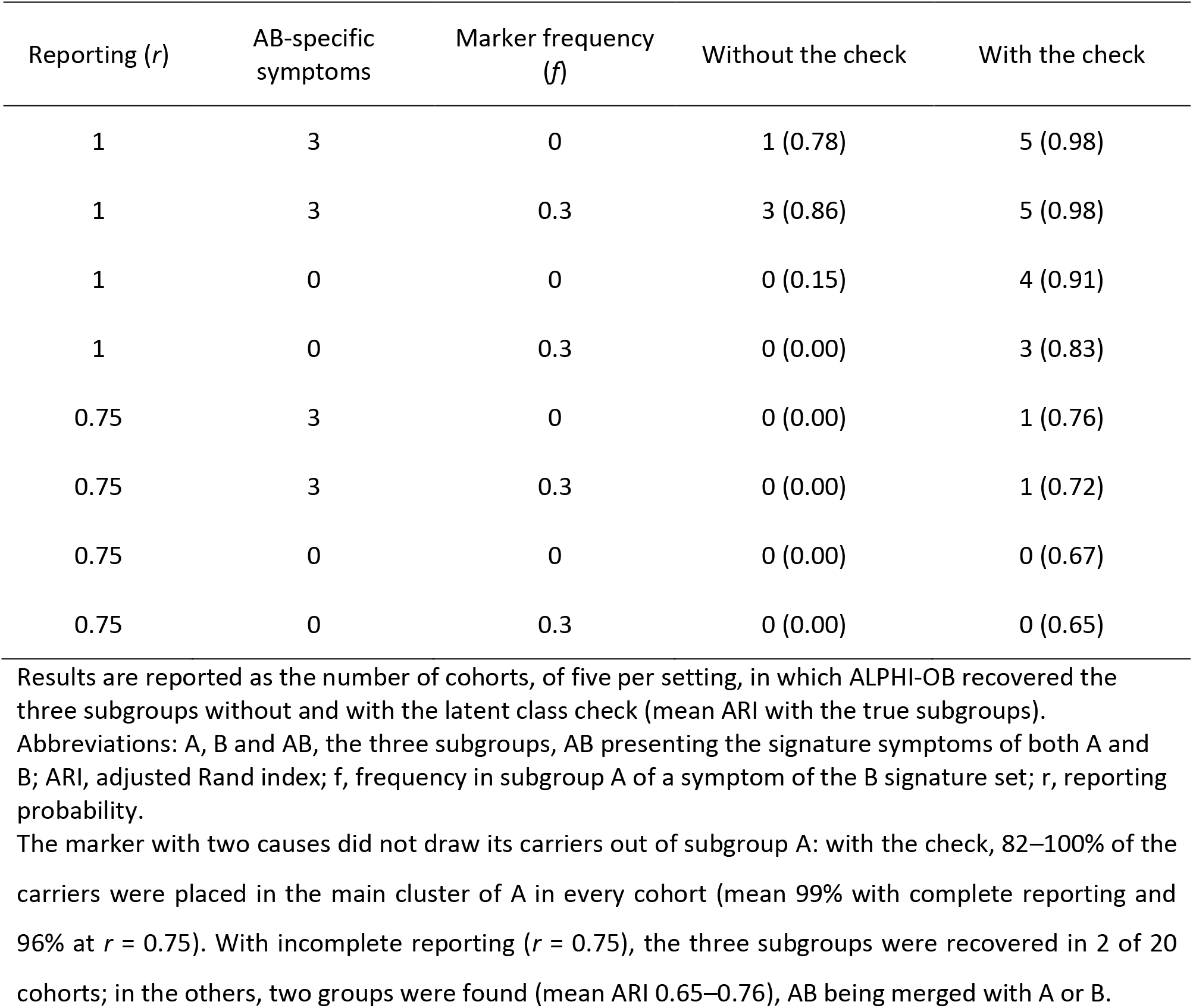
Effect of the latent class check in Simulation 4

The marker with two causes did not draw its carriers out of subgroup A: with the check, 82–100% of the carriers were placed in the main cluster of A in every cohort (mean 99% with complete reporting and 96% at *r* = 0.75). With incomplete reporting (*r* = 0.75), the three subgroups were recovered in 2 of 20 cohorts; in the others, two groups were found (mean ARI 0.65–0.76), AB being merged with A or B.

### Latent subgroups crossing known syndromes

In Simulation 5, ALPHI-OB recovered the latent subgroups and ignored the syndromes (ARI with the latent subgroups 0.94–1.00 in four cohorts, 0.64 in the fifth; ARI with the syndromes 0.36 or less), and the latent class check added the syndrome split within each subgroup. ALPHI (density-based clustering with the latent class check) recovered the four groups in all five cohorts (ARI 0.97–1.00).

### Comparison with language models

In the synthetic cohorts, the number of cohorts resolved was 20/20 for ALPHI and for Claude, 19/20 for ALPHI-Resnik, 18/20 for ALPHI-OB, 13/20 for ALPHI-Jaccard, 11/20 for Grok, 9/20 for ChatGPT, 5/20 for Gemini and 4/20 for DeepSeek (Table 6; Figure 3). Claude resolved every cohort of every setting. (Subgroups were of equal size in Simulation 2, and Claude’s reasoning summaries showed that the last, borderline patients were assigned to balance the group sizes: its accuracy on these cohorts may therefore be overestimated.) ChatGPT and Gemini failed mostly by returning a single group for cohorts with two or three subgroups, and DeepSeek by dividing cohorts into four to eight groups. Gemini returned two groups unrelated to the true subgroups for two D1S2 cohorts (ARI ≈ 0), and Grok frequently added a small extra group to otherwise correct groupings. In the cohorts without subgroups (D1S0), ALPHI, Claude, ChatGPT and Gemini returned a single group for all five cohorts; Grok divided one cohort into four groups, and DeepSeek three cohorts into five to seven groups.

**Figure 3.**
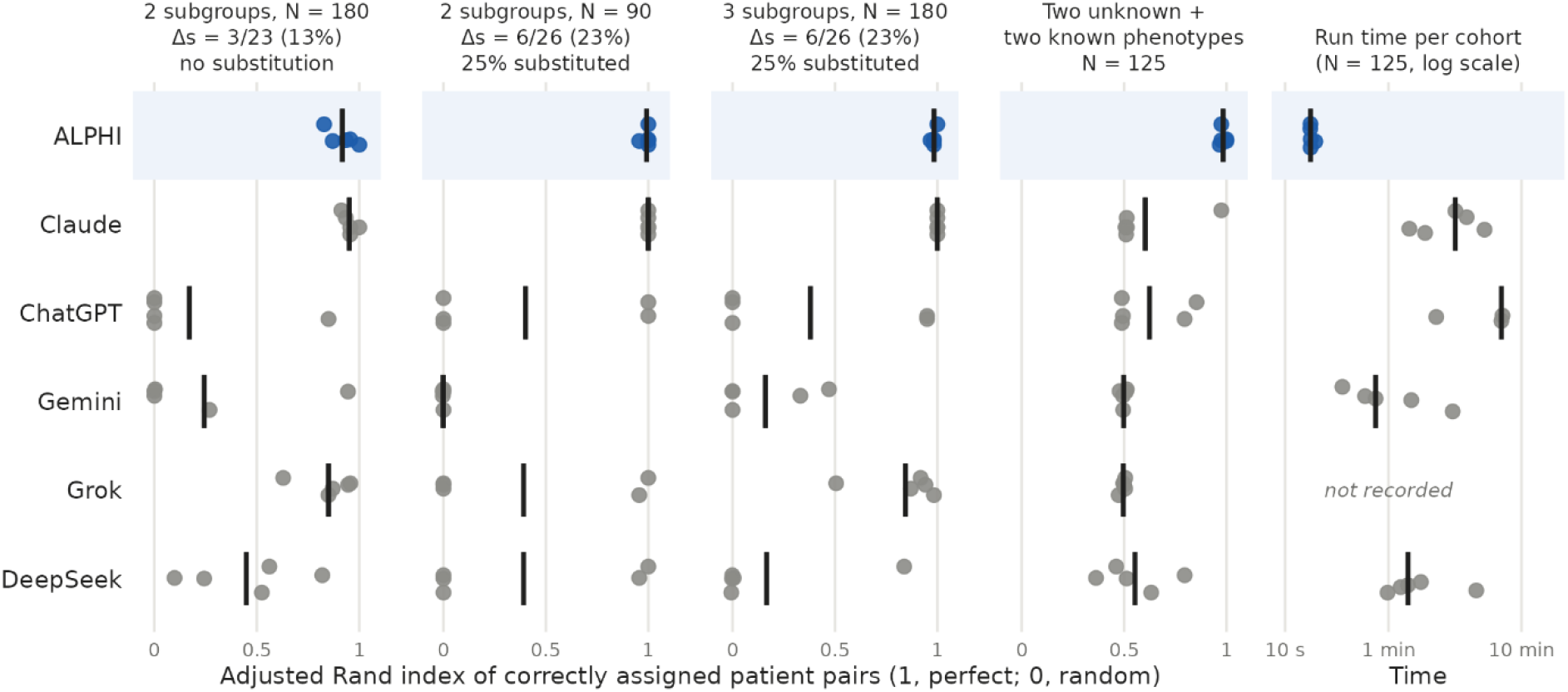
ALPHI and five language models on the same synthetic cohorts. (A) adjusted Rand index between the groups returned and the true subgroups, for three settings of Simulation 2 and for Simulation 5 (two latent subgroups crossing two syndromes, four groups in all). Dots: individual cohorts (five per setting); vertical bars: mean. Δs as in Figure 1. (B) Time per cohort of Simulation 5 (log scale); vertical bars: median. Model times were measured from submission of the prompt to the end of the answer in each provider’s web interface; ALPHI times are wall-clock times on one desktop computer (Intel Core i7-14700). Grok times were not recorded.

**Table 6.** Synthetic cohorts resolved by ALPHI and by five language models

| Method | D1S2,<br>N = 90, Δs = 6,<br>*p* = 0.25 | D1S2,<br>N = 180, Δs = 3,<br>*p* = 0 | D1S3,<br>N = 180, Δs = 6,<br>*p* = 0.25 | D1S0,<br>N = 180, Δs = 6,<br>*p* = 0.25 | Total |
|---|---|---|---|---|---|
| ALPHI | 5 (0.99) | 5 (0.92) | 5 (0.98) | 5 | 20/20 |
| ALPHI-OB | 5 (0.99) | 3 (0.54) | 5 (0.98) | 5 | 18/20 |
| ALPHI-Jaccard | 1 (0.20) | 5 (0.96) | 2 (0.38) | 5 | 13/20 |
| ALPHI-Resnik | 5 (0.99) | 5 (0.95) | 4 (0.90) | 5 | 19/20 |
| Claude | 5 (1.00) | 5 (0.95) | 5 (1.00) | 5 | 20/20 |
| ChatGPT | 2 (0.40) | 0 (0.17) | 2 (0.38) | 5 | 9/20 |
| Gemini | 0 (0.00) | 0 (0.24) | 0 (0.16) | 5 | 5/20 |
| Grok | 2 (0.39) | 2 (0.85) | 3 (0.84) | 4 | 11/20 |
| DeepSeek | 2 (0.39) | 0 (0.45) | 0 (0.17) | 2 | 4/20 |

Results are reported as the number of cohorts resolved, of five per setting (mean ARI with the true subgroups). Abbreviations: ARI, adjusted Rand index; D1S0, D1S2 and D1S3, one disease with no, two or three latent subgroups; N, cohort size; p, term substitution probability; Δs, number of distinguishing symptoms.

In Simulation 5, the four groups were only recovered in all five cohorts by ALPHI (Table 7). Claude, Grok and Gemini grouped the patients by syndrome in four of the five cohorts each (ARI with the syndromes 0.91–1.00, with the latent subgroups 0.05 or less), ignoring the latent subgroups entirely. ChatGPT and DeepSeek returned between one and six groups, which corresponded to neither factor in most cohorts; ChatGPT recovered the latent subgroups alone in one cohort, as ALPHI-OB did in four. The reasoning summaries of Claude, ChatGPT and Grok showed that the latent signatures were detected, described as “background” symptom sets or as noise, and set aside in favor of the syndromes; Grok reached the same result with clustering code of its own (k-means and agglomerative clustering of Jaccard distances).

**Table 7.**
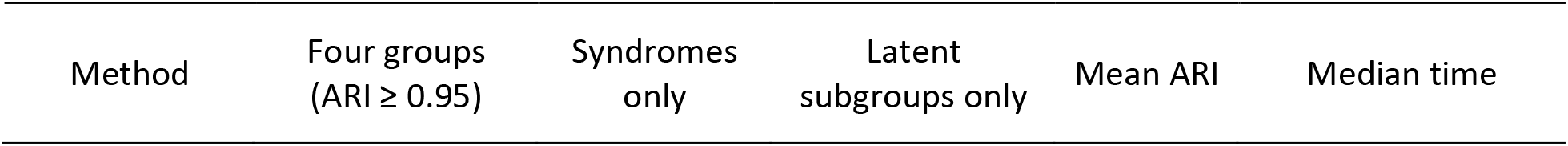
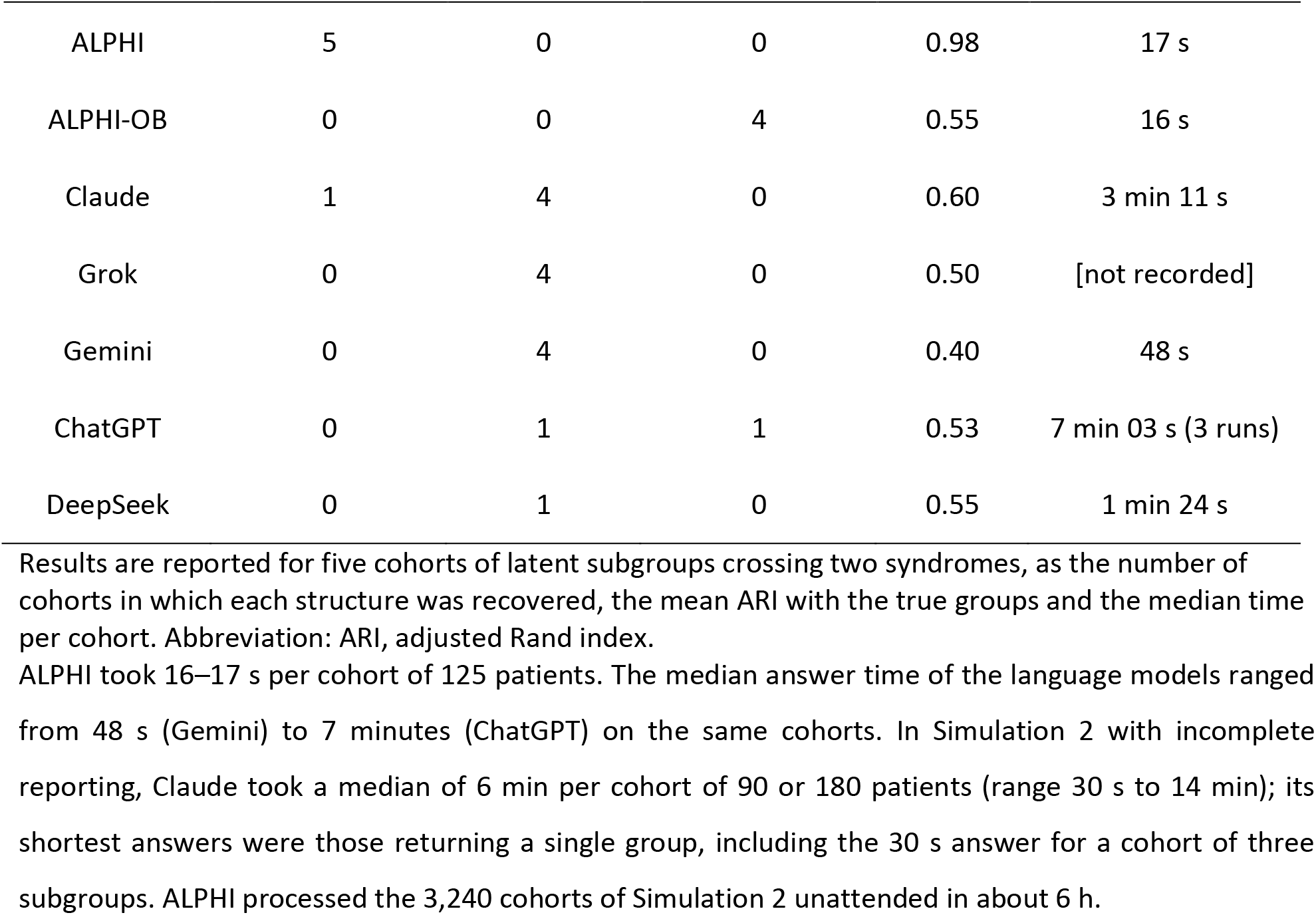
Structure recovered in Simulation 5 by ALPHI and five language models

| Method | Four groups<br>(ARI ≥ 0.95) | Syndromes<br>only | Latent<br>subgroups only | Mean ARI | Median time |
|---|---|---|---|---|---|
| ALPHI | 5 | 0 | 0 | 0.98 | 17 s |
| ALPHI-OB | 0 | 0 | 4 | 0.55 | 16 s |
| Claude | 1 | 4 | 0 | 0.60 | 3 min 11 s |
| Grok | 0 | 4 | 0 | 0.50 | [not recorded] |
| Gemini | 0 | 4 | 0 | 0.40 | 48 s |
| ChatGPT | 0 | 1 | 1 | 0.53 | 7 min 03 s (3 runs) |
| DeepSeek | 0 | 1 | 0 | 0.55 | 1 min 24 s |

Results are reported for five cohorts of latent subgroups crossing two syndromes, as the number of cohorts in which each structure was recovered, the mean ARI with the true groups and the median time per cohort. Abbreviation: ARI, adjusted Rand index.

ALPHI took 16–17 s per cohort of 125 patients. The median answer time of the language models ranged from 48 s (Gemini) to 7 minutes (ChatGPT) on the same cohorts. In Simulation 2 with incomplete reporting, Claude took a median of 6 min per cohort of 90 or 180 patients (range 30 s to 14 min); its shortest answers were those returning a single group, including the 30 s answer for a cohort of three subgroups. ALPHI processed the 3,240 cohorts of Simulation 2 unattended in about 6 h.

### *MYO5B*-related disease

With ALPHI-OB, the 137 *MYO5B* patients were divided into three clusters, with no patient left unassigned (Figure 4). The two-cluster solution obtained a lower selection score (0.78 vs 0.92). All three clusters were stable on resampling (mean Jaccard index 0.99, 0.93 and 0.83), and none was flagged by the sparse-feature check, each pair of clusters differing on 10 to 15 symptoms. The latent class check was concordant with this solution (three classes; ARI 0.92 with the ALPHI-OB clusters; mean resampling Jaccard index 0.95–0.99), and the ALPHI-OB clusters were retained as the ALPHI solution. Five patients of the MVID+PFIC cluster were placed differently by the latent class model, three in the PFIC class and two in the MVID class (Figure 4).

**Figure 4.**
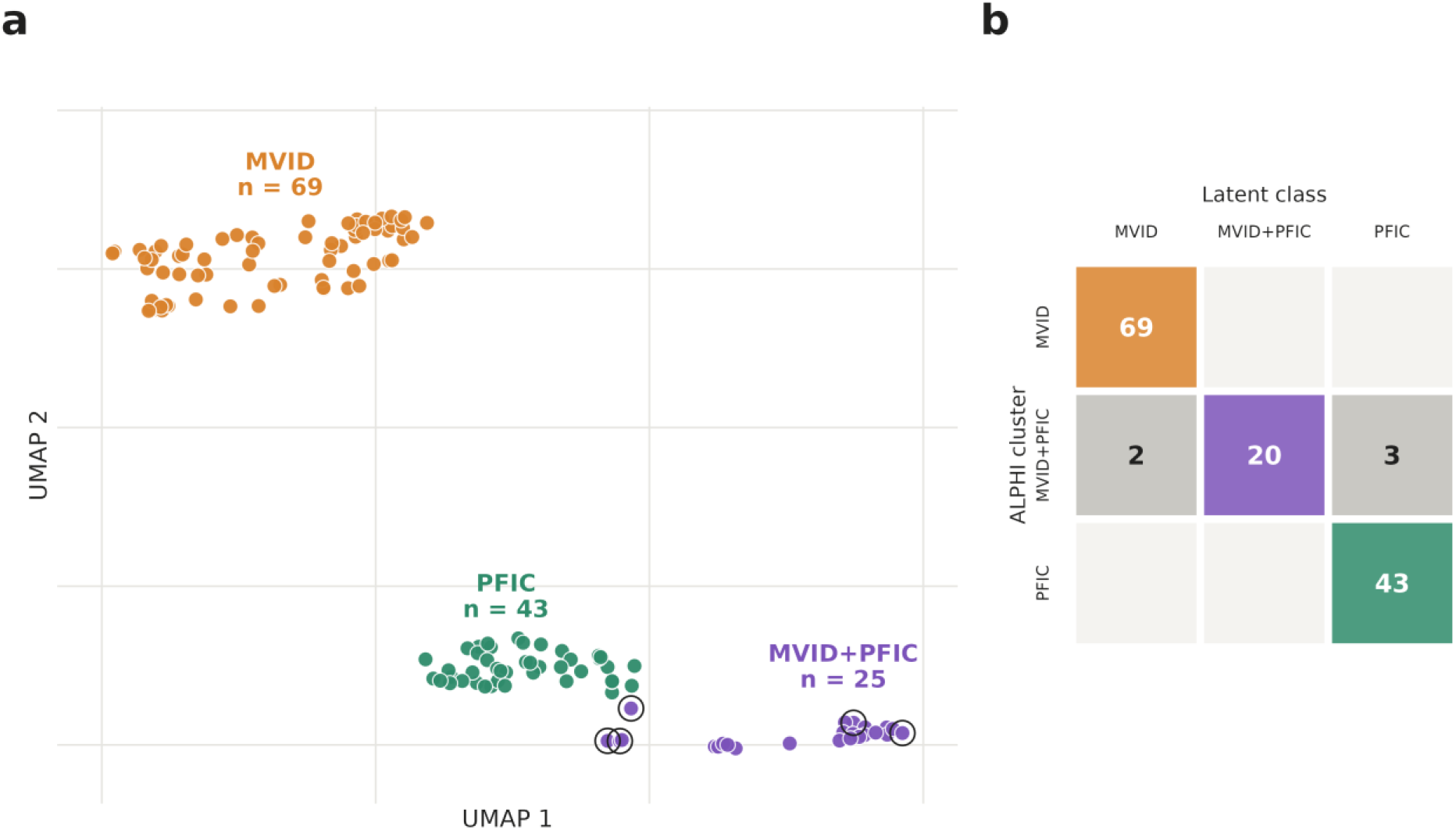
*MYO5B* patients in UMAP space. (A) The 137 patients colored by ALPHI cluster; ringed: the five patients placed in another group by the latent class model of the check. (B) ALPHI clusters against latent classes (numbers of patients; adjusted Rand index 0.92). The two-dimensional embedding is for display; clusters were found in a ten-dimensional embedding of the same distances.

The three clusters corresponded to distinct presentations (Figure 5). The MVID signature was present in 60 of 69 patients of the MVID cluster, 19 of 25 of the MVID+PFIC cluster and none of the PFIC cluster. The PFIC signature was present in 29 of 43, 12 of 25 and none of the MVID cluster, respectively. The MVID+PFIC cluster was the nearest cluster to both others (note that the location of the clusters in the UMAP embedding, Figure 4, is not representation of true inter-cluster distances). Each cluster comprised patients from several articles (19, 11 and 11), and 6 of the 35 articles contributed patients to more than one cluster.

**Figure 5.**
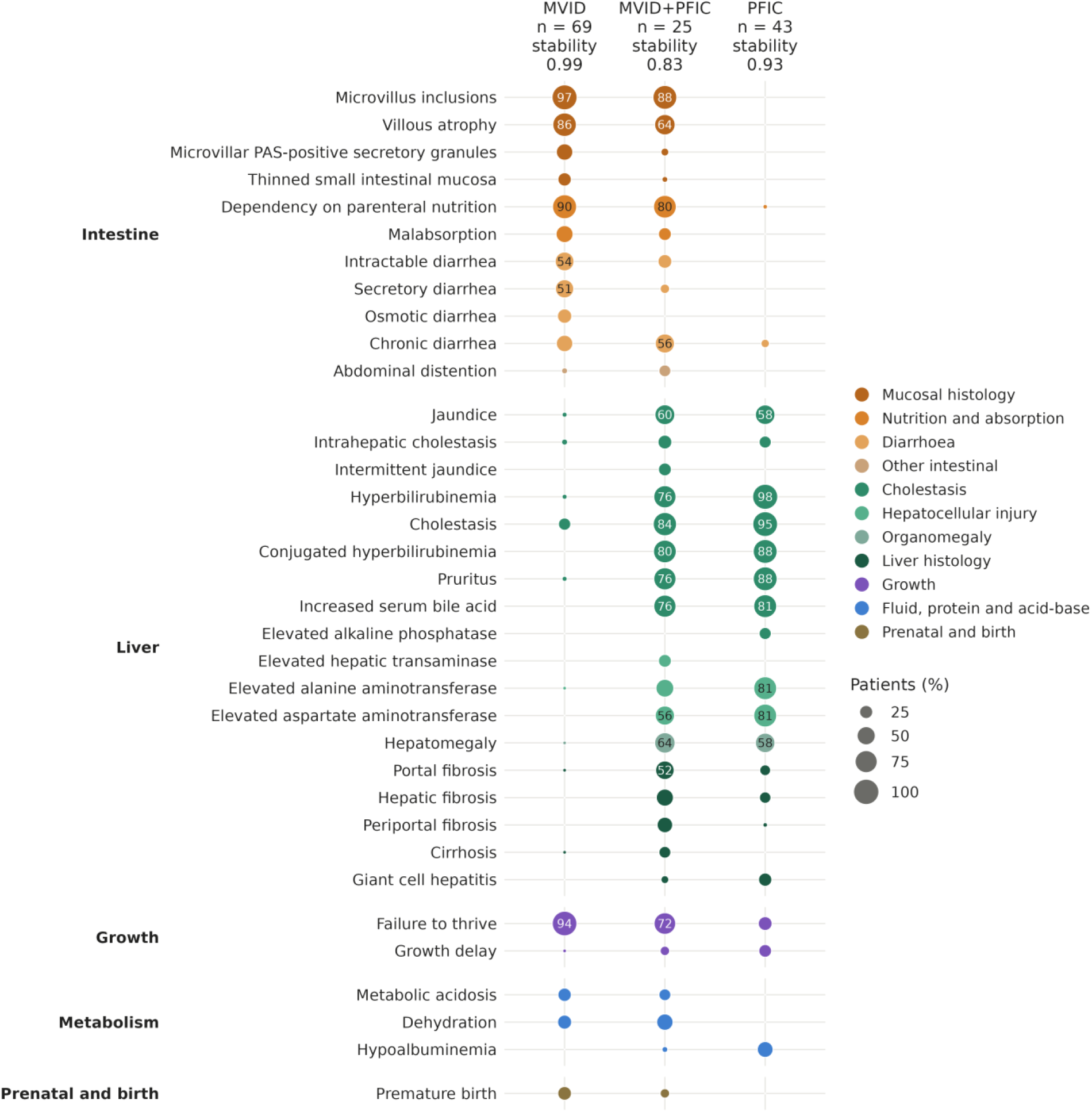
Phenotypic makeup of the *MYO5B* clusters (ALPHI). Prevalence of the HPO terms whose frequency differs by at least 20 percentage points between clusters, grouped by organ and colored by organ subgroup. Dot area and number: percentage of patients. Stability: mean Jaccard index of each cluster over 50 subsamples of 80% of patients. No pair of clusters was flagged by the sparse-feature check.

Symptoms explicitly reported as absent, which were not used for clustering, were consistent with this division (Figure S7). At least one symptom was reported absent in 127 of the 137 patients. Microvillus inclusions and villous atrophy were each reported absent in 6 of 43 patients of the PFIC cluster, but in at most one patient of the other clusters. Conversely, cholestasis was reported absent only in the MVID cluster (8 of 69 patients). Elevated γ-glutamyltransferase was reported absent in 35 of 43 patients of the PFIC cluster (81%) and 17 of 25 of the MVID+PFIC cluster (68%), compared with 11 of 69 of the MVID cluster (16%). Other absences reflected the investigations of each presentation: polyhydramnios (30 of 69) and abnormal renal function (21 of 69) in the MVID cluster, and biliary obstruction (8 of 25) in the MVID+PFIC cluster. No patient of the PFIC or MVID+PFIC cluster was reported free of cholestasis. Premature birth was reported in 19 of 69 patients of the MVID cluster and 3 of 25 of the MVID+PFIC cluster, but in none of the PFIC cluster, in which it was reported absent in 15 of 43 patients.

With ALPHI-Resnik and ALPHI-Jaccard, the optimal solution comprised two clusters, MVID (71 patients) and PFIC (66), the MVID+PFIC patients being included in the PFIC cluster (Figures S8–S13). The three-cluster alternative isolated the MVID+PFIC patients with ALPHI-Resnik (stability 0.86). With ALPHI-Jaccard, it instead divided the MVID patients according to the term used for their diarrhea (chronic vs intractable or secretory); this division was unstable (mean Jaccard index 0.63; dissolution in 54% of subsamples).

### Genotype and survival

Genotypes and survival, which were not used for clustering, differed between the ALPHI clusters (Figure 6). Among patients with both variants classified, biallelic null genotypes were present in 14 of 27 (52%) in the MVID cluster, 5 of 13 (38%) in the MVID+PFIC cluster and 4 of 42 (10%) in the PFIC cluster (permutation test, p < 0.001); 30 of the 42 PFIC patients carried at least one missense variant. Thirty-eight of the 69 MVID patients had not been genetically tested, most of them reported before genetic testing became available (median year of publication 1992, against 2021 for the genotyped MVID patients).

**Figure 6.**
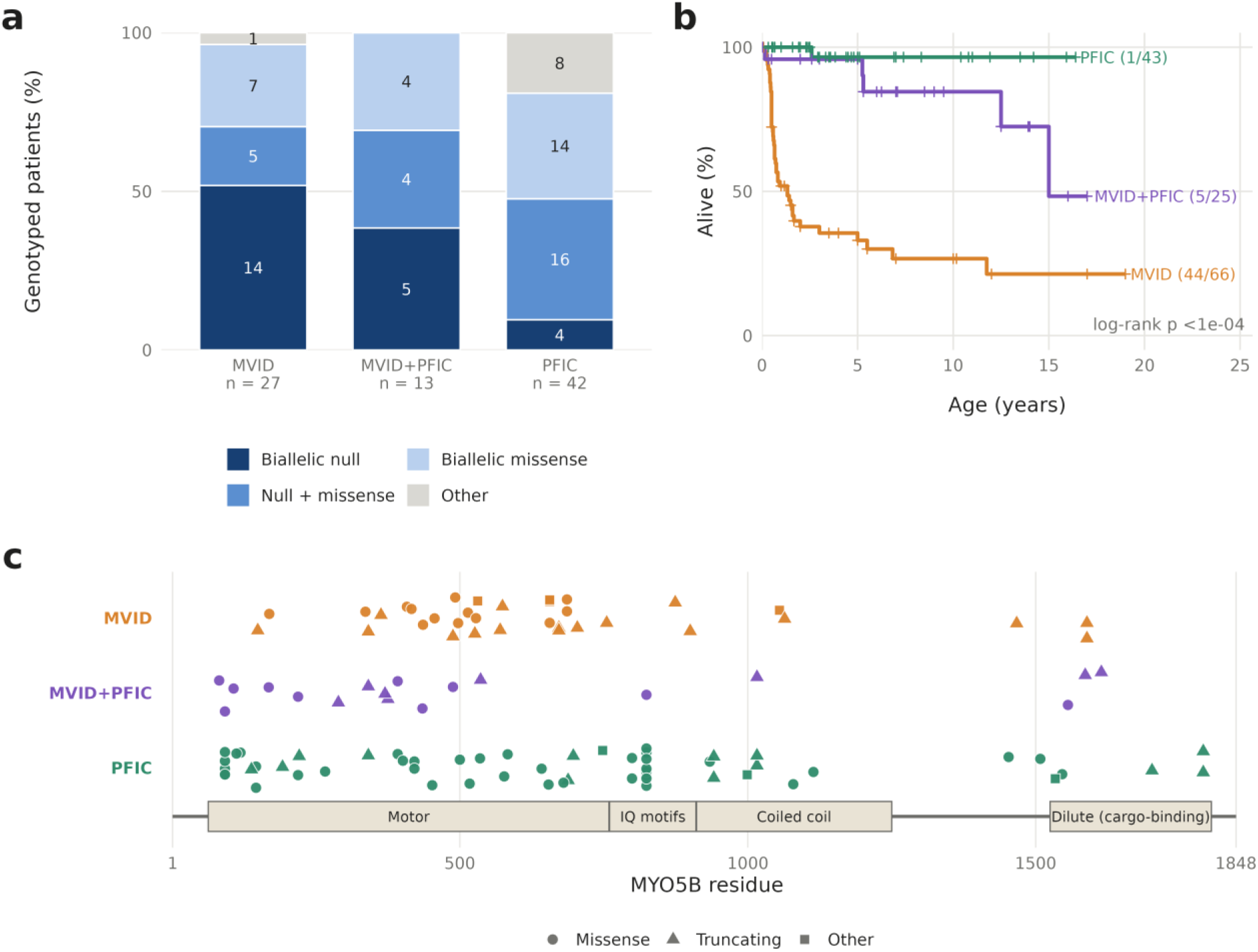
Genotype and survival of the *MYO5B* clusters (ALPHI). Neither was used for clustering. (A) Genotype class of patients with both variants classified (null: nonsense, frameshift or splice site). (B) Kaplan–Meier survival from birth; deaths/patients in brackets; tick marks, censoring at last follow-up. (C) Position of variants on the *MYO5B* protein, one track per cluster; splice variants without a protein position are not shown. Domain boundaries from UniProt Q9ULV0.

Deaths were reported in 44 of 66 patients of the MVID cluster (median age at death 0.6 years), 5 of 25 of the MVID+PFIC cluster and 1 of 43 of the PFIC cluster (log-rank p < 0.0001). The same difference was observed in the subgroup of genotyped patients (19 deaths among 31 MVID patients) and among patients reported from 2010 onwards (18 of 30).

### Term heterogeneity and age at report

In simulated cohorts, the estimated proportion of features coded with a substitute matched the simulated substitution probability (mean 0.003 at *p* = 0, 0.10 at *p* = 0.1, 0.25 at *p* = 0.25, 0.39 at *p* = 0.4 and 0.48 at *p* = 0.5). In the *MYO5B* cohort, it was 0.09 (0.11 in the MVID cluster, 0.03 in the PFIC cluster and 0.11 in the MVID+PFIC cluster), and 0.12 when terms two steps apart were included. Diarrhea accounted for most substitutions (chronic, intractable or secretory diarrhea: 32 of the 61 substitutes in the MVID cluster). Some of the pairs counted were distinct features rather than substitutes (e.g. hepatomegaly and hepatic fibrosis), so that these values are upper estimates. The *MYO5B* cohort therefore lies between *p* = 0 and *p* = 0.25 in the simulations.

Among patients of the MVID and MVID+PFIC clusters, the proportion assigned to the MVID+PFIC cluster increased with age at last report: 4 of 38 (11%) below 1 year, 4 of 25 (16%) between 1 and 5 years, and 17 of 28 (61%) above 5 years (Mann–Whitney test on age, p < 0.001). Onset was neonatal in both clusters (median 0.1 and 0.2 months, compared with 2 months in the PFIC cluster). The difference in age largely reflected the early deaths of MVID patients: among surviving patients, age at last report did not differ significantly between the two clusters (median 3.8 vs 6.6 years; p = 0.23). The number of reported terms was not significantly correlated with age in any cluster.

### Comparison with language models in the *MYO5B* cohort

In the *MYO5B* cohort, the PFIC cluster was recovered by every model: in 67 of 75 runs, one group contained at least 36 of its 43 patients and at most three other patients. The MVID+PFIC cluster was rarely isolated. It formed a group with at most five other patients in 7 of 75 runs (Grok, 4; Claude, 2; Gemini, 1), was merged with the MVID cluster in 9 runs (7 of them by Claude, which then returned only two groups), and was otherwise grouped with up to 22 patients of the MVID cluster; 22 patients of the MVID cluster reported at least one hepatic term. ChatGPT, DeepSeek and Gemini frequently divided the MVID cluster into two to four groups, including, for DeepSeek, a group of 19 or 20 patients described by their renal features. Mean agreement with the ALPHI-OB clusters ranged from 0.44 to 0.75 (Table 8), against 0.17 for a grouping of the patients by article alone. Claude had the highest mean agreement in all three conditions, whereas the run closest to the ALPHI-OB clusters was obtained by Grok (blind condition, ARI 0.95), with the MVID+PFIC patients in a group of their own. Naming the gene or the articles did not increase agreement. For Claude, agreement decreased as information was added (mean ARI 0.75 blind, 0.66 gene, 0.62 article), and with the articles Claude returned two groups in four of five runs; agreement was also highest in the blind condition for Grok, but not for the other models, and the differences between conditions were not significant (Friedman test on the mean ARI of each model, p = 0.25). The mean ARI between runs of the same model and condition ranged from 0.49 to 0.82. In the negative control, Claude, ChatGPT and Grok returned a single group in every run, Gemini divided one run into two groups (135 and 2 patients), and DeepSeek two runs into five and six groups. No answer mentioned an external source, and no disease label was named in the blind and control conditions.

**Table 8.** Groupings of the *MYO5B* cohort by five language models

| Model | Blind | Gene | Article | Groups per run | ARI between runs (blind / gene / article) | Control runs split |
|---|---|---|---|---|---|---|
| Claude | 0.75 (0.61–0.90) | 0.66 (0.61–0.81) | 0.62 (0.61–0.62) | 2–3 | 0.74 / 0.74 / 0.82 | 0/5 |
| ChatGPT | 0.47 (0.42–0.49) | 0.44 (0.37–0.50) | 0.48 (0.37–0.62) | 3–6 | 0.71 / 0.69 / 0.68 | 0/5 |
| Gemini | 0.52 (0.47–0.60) | 0.49 (0.42–0.58) | 0.58 (0.50–0.85) | 2–7 | 0.54 / 0.71 / 0.49 | 1/5 |
| Grok | 0.66 (0.49–0.95) | 0.50 (0.32–0.61) | 0.55 (0.49–0.64) | 2–6 | 0.50 / 0.74 / 0.68 | 0/5 |
| DeepSeek | 0.45 (0.36–0.54) | 0.45 (0.34–0.53) | 0.47 (0.45–0.49) | 3–7 | 0.68 / 0.70 / 0.80 | 2/5 |

Results are reported as mean ARI (five runs per condition) with the ALPHI-OB clusters (range), number of groups, mean ARI between runs, and negative-control runs split. Blind, no gene or disease named; gene, all patients stated to carry pathogenic variants in *MYO5B*; article, gene named and source article of each patient given. Abbreviation: ARI, adjusted Rand index.

## Discussion

The main finding of this study is that phenotypic subgroups can be recovered from literature patient cohorts without recourse to diagnostic labels. In simulated cohorts, spurious subgroups were extremely rare, and errors consisted of merges rather than splits. In simulated cohorts of 44 distinct syndromes, 41 to 44 syndromes were recovered by ALPHI in each cohort. In 137 published patients with *MYO5B*-related disease, MVID, PFIC and a combined MVID+PFIC presentation were separated. These three groups were cross-validated in terms of absent symptoms, genotype and survival. Their recovery from reported phenotypes alone also shows that these presentations exist independently of their clinical interpretation, and that their existence can now be tested objectively.

### Choice of distance metric

No single version was optimal in all situations. ALPHI-Jaccard performed best when the same term was used for the same feature in every report, but failed as soon as related terms were substituted. Literature cohorts correspond to the latter situation: diarrhea for instance, is described as chronic, intractable or secretory depending on the article. In the *MYO5B* cohort, the three-cluster ALPHI-Jaccard solution accordingly divided the MVID patients by the adjective used for their diarrhea. ALPHI-OB, whose distance gives partial credit to neighboring terms within an organ system, was the most reliable version under term substitution and recovered the three *MYO5B* presentations as its optimal solution. In Simulation 1, in which features were reported with exact terms, ALPHI-Jaccard and ALPHI-OB separated the 44 syndromes almost equally well, and both were little affected by random noise terms.

ALPHI-Resnik required the fewest patients to separate two or three distinct diseases, particularly with incomplete reporting, but was the least accurate in separating the 44 syndromes of Simulation 1, especially in the presence of noise. As Resnik similarity is driven by the information content of shared terms, a few specific terms can outweigh many common ones, which favors the separation of diseases that differ by specific features. Conversely, a random noise term can share an informative ancestor with a genuine feature of another disease, which may explain the sensitivity of ALPHI-Resnik to noise. Subgroups within one disease, in contrast, share its core symptoms, against which the distinguishing terms must stand out. Consistent with this, ALPHI-Resnik varied most between compositions in Simulation 2 and identified the MVID+PFIC group only as an alternative solution. ALPHI-OB is therefore recommended for literature cohorts, with ALPHI-Resnik as a complementary analysis.

The 44 syndromes of Simulation 1 were those used to evaluate Phenomizer [3], but the two analyses address different problems. Phenomizer is supervised and works top-down, from known disease phenotypes to the patient: the features of a single patient are compared with reference annotations of known diseases, and these diseases are ranked. ALPHI is unsupervised and works bottom-up, from the patients to the phenome: no disease is defined in advance, and the groups emerge from the similarities between patients alone, with 20 patients per syndrome and about 11 features per patient in Simulation

A supervised approach can only recognize diseases present in its reference annotations; it can indicate that no known disease fits a patient, but cannot identify or describe a disease that has not yet been delineated, whereas an unsupervised approach can, provided that enough patients share the phenotype. The same pattern was nevertheless observed in both studies: randomly added noise terms had little effect, and methods based on exact term matching failed only when terms were imprecise or substituted. Resnik similarity, which placed the correct diagnosis first for most simulated patients with Phenomizer, was the least robust version of ALPHI in the presence of noise. This difference is consistent with the two designs: a patient compared with the full annotation of a disease is matched against many reference terms, whereas two sparse patient profiles share few terms, so that a single spurious match weighs more.

### False positives

A single failure mode of ALPHI-OB was identified in Simulations 2 and 3: in large cohorts with complete reporting, a frequent symptom that is often the only one of its organ system can divide a group in two. Matching such symptoms to the closest symptom anywhere in the other patient makes this event rare without completely abolishing it. Its practical impact is limited by two observations. First, it occurred only with complete reporting and cohorts of 180 patients or more, conditions rarely met by literature cohorts. Second, 11 of the 12 such splits examined were flagged by the sparse-feature check, whereas no true subgroup was flagged. The check is therefore reported with every solution, and a flagged split should not be interpreted without further evidence.

### Comparison with language models

Among the five evaluated LLMs, only Claude approached the accuracy of ALPHI, and even Claude failed on the most demanding simulated cohorts in four of five runs. The four other models resolved 4 to 11 of the 20 synthetic cohorts, and two of them divided cohorts without subgroups. The reliability of a grouping, moreover, cannot be judged from a single answer: the models gave no measure of stability, their groupings of the *MYO5B* cohort varied between runs (mean ARI between runs 0.49–0.82), and the same model returned two groups in one run and three in another. ALPHI gives the same result for a given seed, and reports the stability and sparse-feature checks of every cluster. Furthermore, while ALPHI runs at little or no cost, in seconds per cohort, whereas LLMs are notoriously resource intensive and costly to users and the environment [23]. Lastly, submitting a cohort to a commercial LLM transfers patient data to an external service, contravening data-protection rules, whereas ALPHI (including in its browser version) involves no data transfer whatsoever.

In the *MYO5B* cohort, all models recovered the PFIC presentation, but the MVID+PFIC cluster was isolated in only 7 of 75 runs. In most runs, it was combined with up to 22 patients of the MVID cluster, a number matching that of the MVID patients with a hepatic term. This suggests that the models grouped patients by the presence of any liver involvement, whereas ALPHI-OB retained patients with isolated cholestasis, probably associated with parenteral nutrition, in the MVID cluster. Naming the gene or the articles did not bring the groupings closer to the ALPHI-OB clusters, and Claude diverged increasingly from these clusters, which were supported by genotype and survival, as more information was provided in the prompt (mean ARI 0.75 blind, 0.66 with the gene and 0.62 with the articles); with the articles, Claude returned only two groups in four of five runs. This illustrates an intrinsic limitation of language models in weighing the importance of potentially conflicting pieces of information [24]. The synthetic cohorts, for which no published knowledge exists, show what the models can achieve from symptoms alone; in published cohorts, the use of their knowledge of the literature cannot be excluded, and their groupings cannot be considered agnostic even when no disease is named.

### Minimum data required to define a phenotype

The simulations provide an estimate of the numbers of patients and of reported features required for a phenotype to be identified without prior knowledge. With complete reporting, two distinct diseases were separated by ALPHI from cohorts of 10 patients, in which the smaller group comprised three or four patients, and three diseases from 20–45 patients; with half of the symptoms reported, 10–20 and 45–120 patients were required, respectively. Distinguishing latent subgroups of a single disease required more patients, the more so the fewer distinguishing features were available. A new phenotype therefore becomes identifiable once a few tens of patients have been described individually and in sufficient detail, a number that a single case series rarely reaches: recently described gene–disease associations were defined from a median of three patients, with less than half of the phenome documented [25]. Summary tables reporting the frequency of each feature across a series cannot be used for this purpose, since the co-occurrence of features within individual patients is lost. The publication of individual case reports, including those of patients resembling patients already described, is therefore not redundant: each report adds to the number of patients on which the delineation of a disease and of its subtypes depends [25–29]. Conversely, the absence of distinct clusters in a small cohort does not establish that a disease is homogeneous; it may only indicate that too few patients have been described.

#### *MYO5B*-related disease

The three ALPHI-OB clusters reproduce the classification of *MYO5B*-related disease into MVID, PFIC and combined forms proposed by Aldrian et al. [10], but were derived from reported phenotypes alone. The genotype distribution is also consistent with that study: biallelic null genotypes predominated in the MVID cluster, whereas most patients of the PFIC cluster carried at least one missense variant, compatible with a residual protein function that spares the intestine. The MVID+PFIC cluster was intermediate in both genotype and survival.

Cholestasis was mentioned in 22% of the MVID cluster without hyperbilirubinemia, pruritus or increased bile acids. This probably reflects cholestasis associated with parenteral nutrition [30] rather than *MYO5B*-related liver disease, and these patients were retained in the MVID cluster by ALPHI-OB. In the PFIC and MVID+PFIC clusters, γ-glutamyltransferase was explicitly reported as normal in most patients (81% and 68%), in keeping with the low-GGT cholestasis described in *MYO5B* deficiency [31, 32].

### Limitations

The simulations were simplified: symptoms were generated independently of one another, at frequencies derived from Orphanet classes or from Köhler et al. [3], and substitution was limited to terms at the same level or one level up in the HPO graph; the imprecision step of Köhler et al. [3], in which every term is replaced by an ancestor, was not reproduced in Simulation 1. Real cohorts include correlated symptoms, article-dependent reporting and coding errors. In the *MYO5B* cohort, most of the 35 articles contributed patients to a single cluster, so that the reporting style of an article and the phenotype of its patients cannot be fully dissociated. Forty patients, 38 of the MVID cluster and 2 of the MVID+PFIC cluster, had not been genetically tested and were attributed to *MYO5B* on clinical grounds; some may have other causes of MVID. These patients were nevertheless assigned to clusters together with genotyped patients and did not form a separate cluster. The threshold of eight terms excluded 73 of 210 patients and favored well-documented patients. Finally, genotype classes were coarse, and splice variants could not be positioned on the protein.

Lastly, published reports describe each patient at a single time point, at different ages and stages of disease. Phenotypes may converge as diseases progress and treatment complications add shared features to patients with different conditions. For example, liver diseases of different causes tend toward fibrosis and cirrhosis, long-term parenteral nutrition can lead to cholestasis, and central line associated bloodstream infections can lead to neurologic, renal and respiratory impairment. Clusters derived from cross-sectional data may therefore reflect disease stage and treatments as well as the underlying cause.

## Conclusions

Clinically meaningful subgroups can be recovered from patient cohorts without recourse to diagnostic labels. ALPHI recovered the MVID, PFIC and MVID+PFIC presentations of *MYO5B* deficiency and separated 44 distinct syndromes in a single simulated cohort, with a false positive rate of 0.06% (1/1,810). Unlike supervised approaches, which can only recognize diseases already described, ALPHI can also reveal phenotypes that have not yet been delineated. The approach is applicable to any gene or disease for which a sufficient number of patients has been described. ALPHI is freely available as open-source software and as a browser application in which patient data are processed locally.

## Data Availability

The datasets supporting the conclusions of this article are available in the PytheasDB database, https://pytheasdb.com, and in the Zenodo repositories: https://doi.org/10.5281/zenodo.24314916, for the patient data export, software and scripts, and https://doi.org/10.5281/zenodo.24316585 for the simulation results and language model prompts. Simulated cohorts can be regenerated with the scripts provided in the repository.
Project name: ALPHI (agnostic latent phenotype identification)
Project home page: https://github.com/PaulGuerry/ALPHI; browser version at https://alphi.pytheasdb.com
Archived version: https://doi.org/10.5281/zenodo.24314916
Operating system(s): Platform independent
Programming language: Python 3.14; R 4.5.2 (figures)
Other requirements: umap-learn 0.5.12, hdbscan 0.8.44 and the other Python and R packages listed in the repository
License: MIT
Any restrictions to use by non-academics: None

https://doi.org/10.5281/zenodo.24314916

https://pytheasdb.com

https://github.com/PaulGuerry/ALPHI

https://alphi.pytheasdb.com

## Supplementary Information

Supplementary Information (PDF): supplementary figures and tables. Figures S1–S13: recovery of latent subgroups and separation of distinct diseases in Simulations 2 and 3 by the three distances without the latent class check (Figures S1–S3); effect of the latent class check (Figures S4–S6); symptoms reported absent in the three MYO5B clusters (Figure S7); UMAP embedding, phenotypic makeup, genotype and survival of the MYO5B clusters obtained with ALPHI-Resnik and ALPHI-Jaccard (Figures S8–S13). Tables S1–S3: Orphanet diseases from which the symptoms of Simulation 2 were drawn (Table S1); recovery of each of the 44 syndromes of Simulation 1 (Table S2); latent class analysis alone in Simulation 1 (Table S3).

## Abbreviations

ALPHI: agnostic latent phenotype identification
ARI: adjusted Rand index
BIC: Bayesian information criterion
DBCV: density-based cluster validity
GGT: gamma-glutamyl transferase
HDBSCAN: hierarchical density-based spatial clustering of applications with noise
HPO: Human Phenotype Ontology
IC: information content
LLM: large language model
MVID: microvillus inclusion disease
OB: organ branch
PFIC: progressive familial intrahepatic cholestasis
SD: standard deviation
UMAP: uniform manifold approximation and projection

## Declarations

### Ethics approval and consent to participate

Only de-identified data on patients described in previously published reports were analyzed; no new patients were recruited. Approval by an ethics committee and informed consent were therefore not required.

### Consent for publication

Not applicable.

### Availability of data and materials

The datasets supporting the conclusions of this article are available in the PytheasDB database, https://pytheasdb.com [17], and in the Zenodo repositories: https://doi.org/10.5281/zenodo.24314916 for the patient data export, software and scripts [20], and https://doi.org/10.5281/zenodo.24316585 for the simulation results and language model prompts [21]. Simulated cohorts can be regenerated with the scripts provided in the repository. Project name: ALPHI (agnostic latent phenotype identification) Project home page: https://github.com/PaulGuerry/ALPHI; browser version at https://alphi.pytheasdb.com Archived version: https://doi.org/10.5281/zenodo.24314916 Operating system(s): Platform independent Programming language: Python 3.14; R 4.5.2 (figures) Other requirements: umap-learn 0.5.12, hdbscan 0.8.44 and the other Python and R packages listed in the repository License: MIT Any restrictions to use by non-academics: None

## Authors’ contributions

Conceptualization: PG, AF; Methodology: PG; Software: PG; Data curation: PG; Formal analysis: PG; Investigation: PG; Visualization: PG; Funding acquisition: AF; Supervision: AF, PG; Writing – original draft: PG; Writing – review & editing: PG, AF. Both authors read and approved the final manuscript.

## Declaration of generative AI use

During the preparation of this work the authors used Claude (Anthropic) to assist with coding and with the editing and formatting of the manuscript. After using this tool, the authors reviewed and edited the content as needed and take full responsibility for the content of the article.

