## Supplementary Information for "Defining diseases from the bottom up with agnostic latent phenotype identification"

### Supplementary tables

**Table S1.** Orphanet diseases from which the symptoms of Simulation 2 were drawn

| Disease | ORPHA | Symptoms in Orphanet list | C1 | C2 | C3 |
|---|---|---|---|---|---|
| Alagille syndrome | 52 | 43 | 7 | 2 | 8 |
| Alport syndrome | 63 | 34 | 4 | 3 | 3 |
| Bardet–Biedl syndrome | 110 | 95 | 11 | 8 | 8 |
| Beckwith–Wiedemann syndrome | 116 | 85 | 6 | 9 | 7 |
| Cystic fibrosis | 586 | 35 | 2 | 3 | 2 |
| Dubowitz syndrome | 235 | 102 | 10 | 13 | 11 |
| Fabry disease | 324 | 76 | 3 | 3 | 12 |
| Glutaryl-CoA dehydrogenase deficiency | 25 | 43 | 6 | 1 | 5 |
| Marfan syndrome | 558 | 68 | 11 | 13 | 4 |
| Neurofibromatosis type 1 | 636 | 77 | 6 | 7 | 8 |
| Noonan syndrome | 648 | 70 | 9 | 9 | 7 |
| Tuberous sclerosis complex | 805 | 62 | 5 | 4 | 6 |
| Williams syndrome | 904 | 186 | 20 | 25 | 23 |
| Wilson disease | 905 | 56 | 11 | 2 | 3 |

Results are reported as the number of symptoms in each Orphanet list and the number of symptoms used in each composition (core and signature symptoms). A symptom listed for several diseases is counted for each of them; no symptom was used in more than one composition. Abbreviations: C1–C3, the three compositions of Simulation 2; ORPHA, Orphanet identifier.

**Table S2.** Recovery of each of the 44 syndromes of Simulation 1

| Syndrome | MIM | Features listed | Features per patient | ALPHI-OB, q = 0 | ALPHI-Jaccard, q = 0 | ALPHI-Resnik, q = 0 | ALPHI-OB, q = 0.5 | ALPHI-Jaccard, q = 0.5 | ALPHI-Resnik, q = 0.5 |
|---|---|---|---|---|---|---|---|---|---|
| Holt–Oram syndrome | 142900 | 9 | 2.0 | 9 | 10 | 6 | 7 | 10 | 4 |
| von Hippel–Lindau syndrome | 193300 | 8 | 2.7 | 10 | 10 | 10 | 10 | 10 | 7 |
| Split-hand/foot malformation with long bone deficiency 1 (SHFLD1) | 119100 | 21 | 2.8 | 10 | 7 | 4 | 10 | 7 | 4 |
| Townes–Brocks syndrome | 107480 | 12 | 3.4 | 10 | 10 | 7 | 9 | 10 | 6 |
| Currarino syndrome | 176450 | 13 | 3.4 | 10 | 10 | 10 | 9 | 10 | 10 |
| Distal arthrogryposis syndrome type 1 | 108120 | 15 | 3.8 | 9 | 10 | 10 | 8 | 10 | 6 |
| Distichiasis–lymphedema syndrome | 153400 | 20 | 3.9 | 10 | 10 | 10 | 10 | 10 | 10 |
| Popliteal pterygium syndrome | 119500 | 13 | 4.6 | 10 | 10 | 10 | 10 | 9 | 9 |
| Neurofibromatosis type 2 | 101000 | 15 | 4.8 | 9 | 9 | 10 | 8 | 9 | 10 |
| Neurofibromatosis type 1 | 162200 | 20 | 5.3 | 9 | 9 | 10 | 8 | 9 | 7 |
| Branchiootorenal syndrome 1 | 113650 | 23 | 5.3 | 10 | 10 | 10 | 10 | 10 | 10 |
| Catel–Manzke syndrome 1 | 302380 | 26 | 6.7 | 10 | 10 | 10 | 10 | 10 | 10 |

| Syndrome | MIM | Features listed | Features per patient | ALPHI-OB, q = 0 | ALPHI-Jaccard, q = 0 | ALPHI-Resnik, q = 0 | ALPHI-OB, q = 0.5 | ALPHI-Jaccard, q = 0.5 | ALPHI-Resnik, q = 0.5 |
|---|---|---|---|---|---|---|---|---|---|
| Camurati–Engelmann disease | 131300 | 13 | 6.8 | 10 | 10 | 10 | 10 | 10 | 10 |
| CHARGE syndrome | 214800 | 29 | 7.0 | 10 | 10 | 10 | 10 | 10 | 10 |
| Nail–patella syndrome | 161200 | 17 | 7.1 | 10 | 10 | 10 | 10 | 10 | 10 |
| Fraser syndrome | 219000 | 51 | 7.4 | 9 | 10 | 10 | 10 | 9 | 10 |
| Treacher Collins syndrome | 154500 | 26 | 7.4 | 10 | 10 | 10 | 10 | 10 | 10 |
| Greig cephalopolysyndactyly syndrome | 175700 | 28 | 7.5 | 10 | 10 | 10 | 10 | 10 | 10 |
| Congenital contractural arachnodactyly (Beals–Hecht syndrome) | 121050 | 24 | 7.5 | 9 | 10 | 10 | 9 | 10 | 8 |
| Ectrodactyly, ectodermal dysplasia, and cleft lip/palate syndrome (EEC1) | 129900 | 40 | 9.7 | 10 | 8 | 7 | 10 | 9 | 9 |
| Kyphomelic dysplasia | 211350 | 17 | 9.8 | 9 | 10 | 9 | 10 | 10 | 10 |
| Tuberous sclerosis | 191100 | 19 | 10.2 | 10 | 10 | 10 | 10 | 10 | 7 |
| Walker–Warburg syndrome | 236670 | 24 | 10.6 | 10 | 10 | 10 | 10 | 10 | 10 |
| Angelman syndrome | 105830 | 23 | 11.0 | 10 | 10 | 10 | 10 | 10 | 10 |
| Bardet–Biedl syndrome | 209900 | 35 | 11.6 | 10 | 10 | 10 | 10 | 10 | 10 |

| Syndrome | MIM | Features listed | Features per patient | ALPHI-OB, q = 0 | ALPHI-Jaccard, q = 0 | ALPHI-Resnik, q = 0 | ALPHI-OB, q = 0.5 | ALPHI-Jaccard, q = 0.5 | ALPHI-Resnik, q = 0.5 |
|---|---|---|---|---|---|---|---|---|---|
| Kabuki syndrome | 147920 | 26 | 11.8 | 10 | 10 | 10 | 10 | 10 | 10 |
| Mowat–Wilson syndrome | 235730 | 23 | 11.8 | 10 | 10 | 10 | 10 | 10 | 10 |
| Baller–Gerold syndrome | 218600 | 43 | 11.9 | 10 | 10 | 10 | 10 | 10 | 10 |
| Aicardi–Goutières syndrome | — | 21 | 12.3 | 10 | 10 | 10 | 10 | 10 | 10 |
| Freeman–Sheldon syndrome | 193700 | 38 | 12.6 | 10 | 10 | 10 | 10 | 10 | 6 |
| Johanson–Blizzard syndrome | 243800 | 32 | 13.4 | 10 | 10 | 10 | 10 | 10 | 10 |
| Cohen syndrome | 216550 | 25 | 13.5 | 10 | 10 | 10 | 10 | 10 | 10 |
| Hydrolethalus syndrome 1 | 236680 | 23 | 14.0 | 10 | 10 | 10 | 10 | 10 | 10 |
| Smith–Lemli–Opitz syndrome | 270400 | 32 | 14.8 | 10 | 10 | 10 | 10 | 10 | 10 |
| Coffin–Siris syndrome | 135900 | 22 | 15.7 | 10 | 10 | 10 | 10 | 10 | 10 |
| Lacrimoauriculodentodigital syndrome type 1 | 149730 | 27 | 16.0 | 10 | 10 | 8 | 10 | 10 | 7 |
| Faciogenital dysplasia (Aarskog–Scott syndrome) | 305400 | 24 | 16.2 | 10 | 10 | 10 | 10 | 10 | 10 |
| Branchiooculofacial syndrome | 113620 | 48 | 17.8 | 10 | 10 | 10 | 10 | 10 | 10 |

| Syndrome | MIM | Features listed | Features per patient | ALPHI-OB, q = 0 | ALPHI-Jaccard, q = 0 | ALPHI-Resnik, q = 0 | ALPHI-OB, q = 0.5 | ALPHI-Jaccard, q = 0.5 | ALPHI-Resnik, q = 0.5 |
|---|---|---|---|---|---|---|---|---|---|
| Coffin–Lowry syndrome | 303600 | 20 | 17.9 | 10 | 10 | 10 | 10 | 10 | 10 |
| Multiple pterygium syndrome, Escobar variant | 265000 | 31 | 19.0 | 10 | 10 | 10 | 10 | 10 | 10 |
| Shprintzen–Goldberg craniosynostosis syndrome | 182212 | 48 | 19.4 | 10 | 10 | 10 | 10 | 10 | 9 |
| Opitz–Kaveggia syndrome | 305450 | 48 | 21.6 | 10 | 10 | 10 | 10 | 10 | 10 |
| Rubinstein–Taybi syndrome | 180849 | 55 | 24.2 | 10 | 10 | 10 | 10 | 10 | 10 |
| Cornelia de Lange syndrome 1 | 122470 | 59 | 27.8 | 10 | 10 | 10 | 10 | 10 | 10 |

Results are reported as the number of cohorts, of 10 per setting, in which the syndrome was recovered (one cluster containing at least half of its patients, the syndrome forming a majority of that cluster), by version of ALPHI and noise level (q). Features listed: features of the syndrome in Köhler et al. [1]; features per patient: mean number of features of the simulated patients, without noise. Syndromes are sorted by features per patient. Abbreviations: MIM, Mendelian Inheritance in Man number; q, noise level (unrelated terms added per patient, as a proportion of its features).

**Table S3.** Latent class analysis alone in Simulation 1

| q | Cohort | Feature families | Lowest BIC: classes (BIC) | ARI | 44 classes from the syndromes: BIC (ARI) |
|---|---|---|---|---|---|
| 0 | 1 | 110 | 7 (38,958) | 0.09 | 58,119 (0.23) |
| 0 | 2 | 109 | 10 (38,604) | 0.16 | 57,712 (0.37) |
| 0 | 3 | 114 | 6 (39,830) | 0.09 | 59,868 (0.24) |
| 0.5 | 1 | 118 | 9 (41,358) | 0.10 | 62,566 (0.29) |
| 0.5 | 2 | 118 | 7 (41,309) | 0.12 | 62,258 (0.25) |
| 0.5 | 3 | 120 | 8 (41,406) | 0.12 | 63,202 (0.19) |

Latent class models with 1–50 classes were fitted to six cohorts of 44 syndromes of 20 patients (the first three of each noise level, q), using the feature families of the latent class check (present in at least 27 patients). Lowest BIC: number of classes with the lowest BIC, and that BIC; ARI: agreement of the corresponding classes with the syndromes; 44 classes from the syndromes: BIC of the 44-class model started from the true syndromes, and ARI of its classes with the syndromes after fitting. Abbreviations: ARI, adjusted Rand index; BIC, Bayesian information criterion; q, noise level.

### Supplementary figures

ALPHI-OB without the latent class check gave the same three *MYO5B* clusters as ALPHI (Figures 4–6), so that its figures are not repeated.

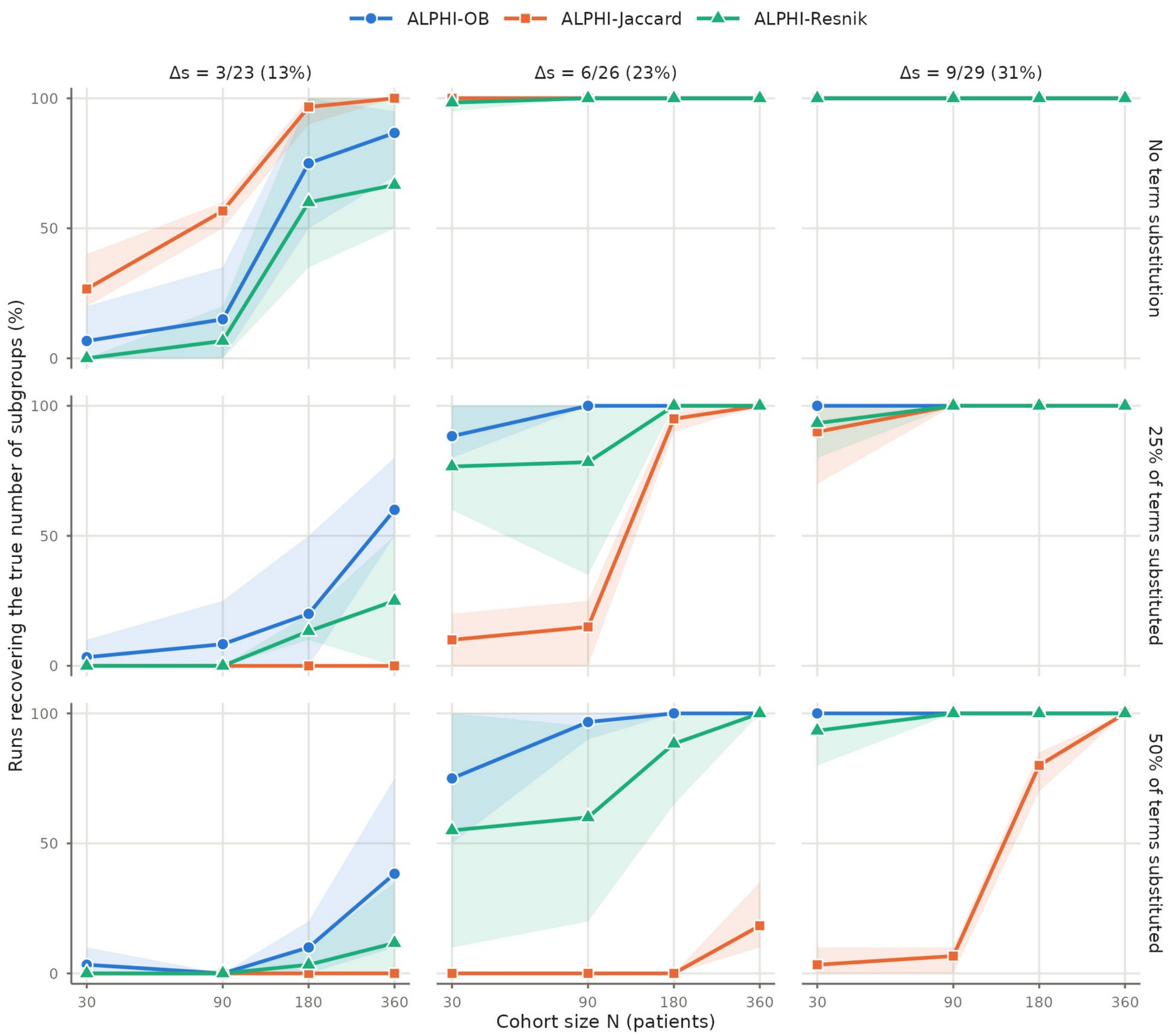


**Figure S1.** Recovery of two latent subgroups (D1S2) in Simulation 2 by the three distances, without the latent class check. Percentage of cohorts in which the true number of subgroups was found by ALPHI-OB (blue circles), ALPHI-Jaccard (orange squares) and ALPHI-Resnik (green triangles), by cohort size N, distinguishing symptoms (Δs, columns) and term substitution (none, 25% or 50% of terms substituted; rows). Δs: number of distinguishing symptoms of a subgroup over the number of symptoms defining it (20 core symptoms shared by all subgroups plus the distinguishing symptoms), with the percentage in parentheses. Lines: mean over compositions C1–C3; shaded bands: range over compositions. Dispersed and concentrated layouts are pooled; complete reporting. Cohorts without subgroups (D1S0) were divided in 1 of 1,08s0 runs with ALPHI-OB, and in none of 1,080 with ALPHI-Jaccard or ALPHI-Resnik.

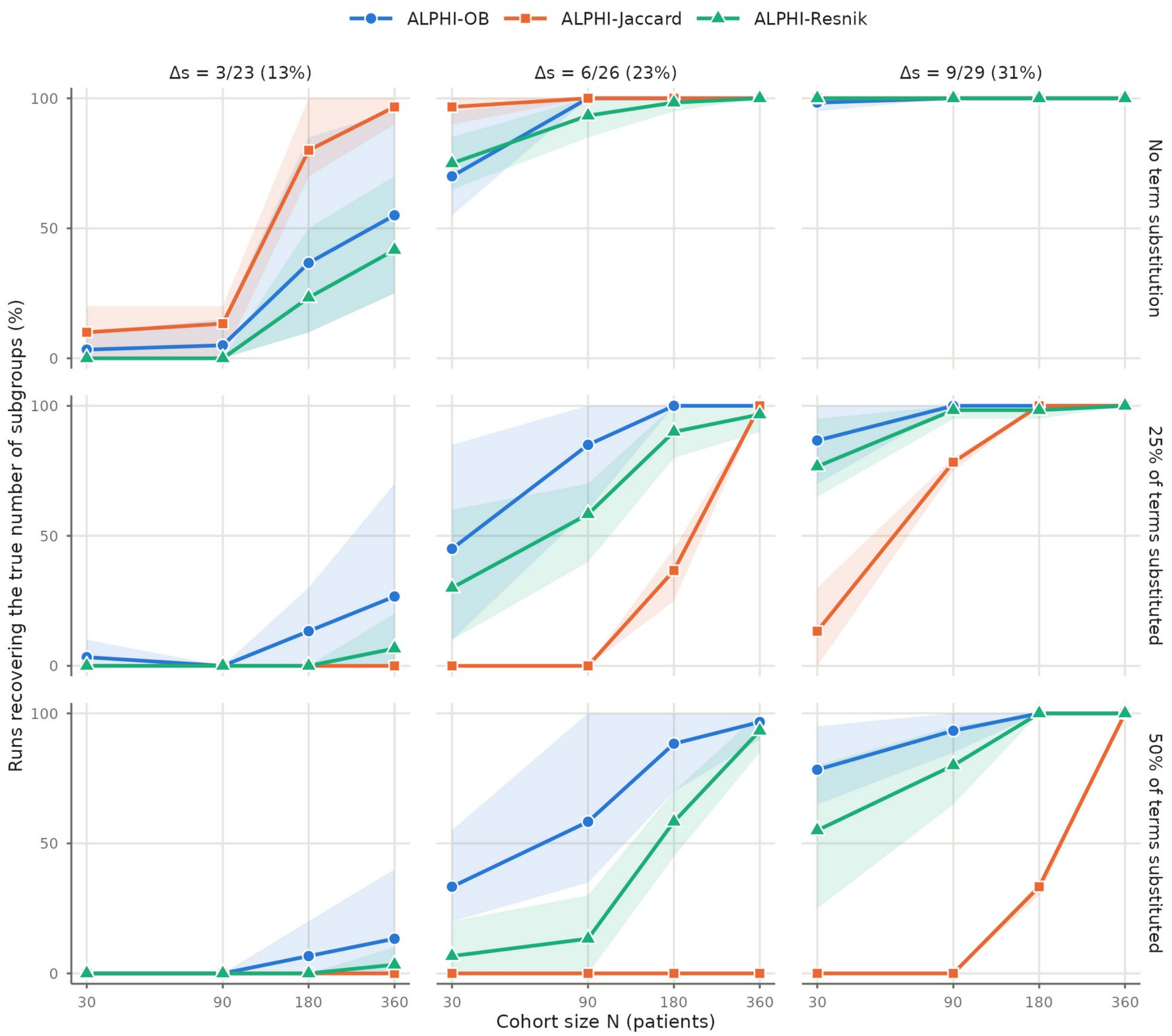


**Figure S2.** Recovery of three latent subgroups (D1S3) in Simulation 2 by the three distances, without the latent class check. Percentage of cohorts in which the true number of subgroups was found by ALPHI-OB (blue circles), ALPHI-Jaccard (orange squares) and ALPHI-Resnik (green triangles), by cohort size N, distinguishing symptoms (Δs, columns) and term substitution (none, 25% or 50% of terms substituted; rows). Δs: number of distinguishing symptoms of a subgroup over the number of symptoms defining it (20 core symptoms shared by all subgroups plus the distinguishing symptoms), with the percentage in parentheses. Lines: mean over compositions C1–C3; shaded bands: range over compositions. Dispersed and concentrated layouts are pooled; complete reporting. Cohorts without subgroups (D1S0) were divided in 1 of 1,080 runs with ALPHI-OB, and in none of 1,080 with ALPHI-Jaccard or ALPHI-Resnik.

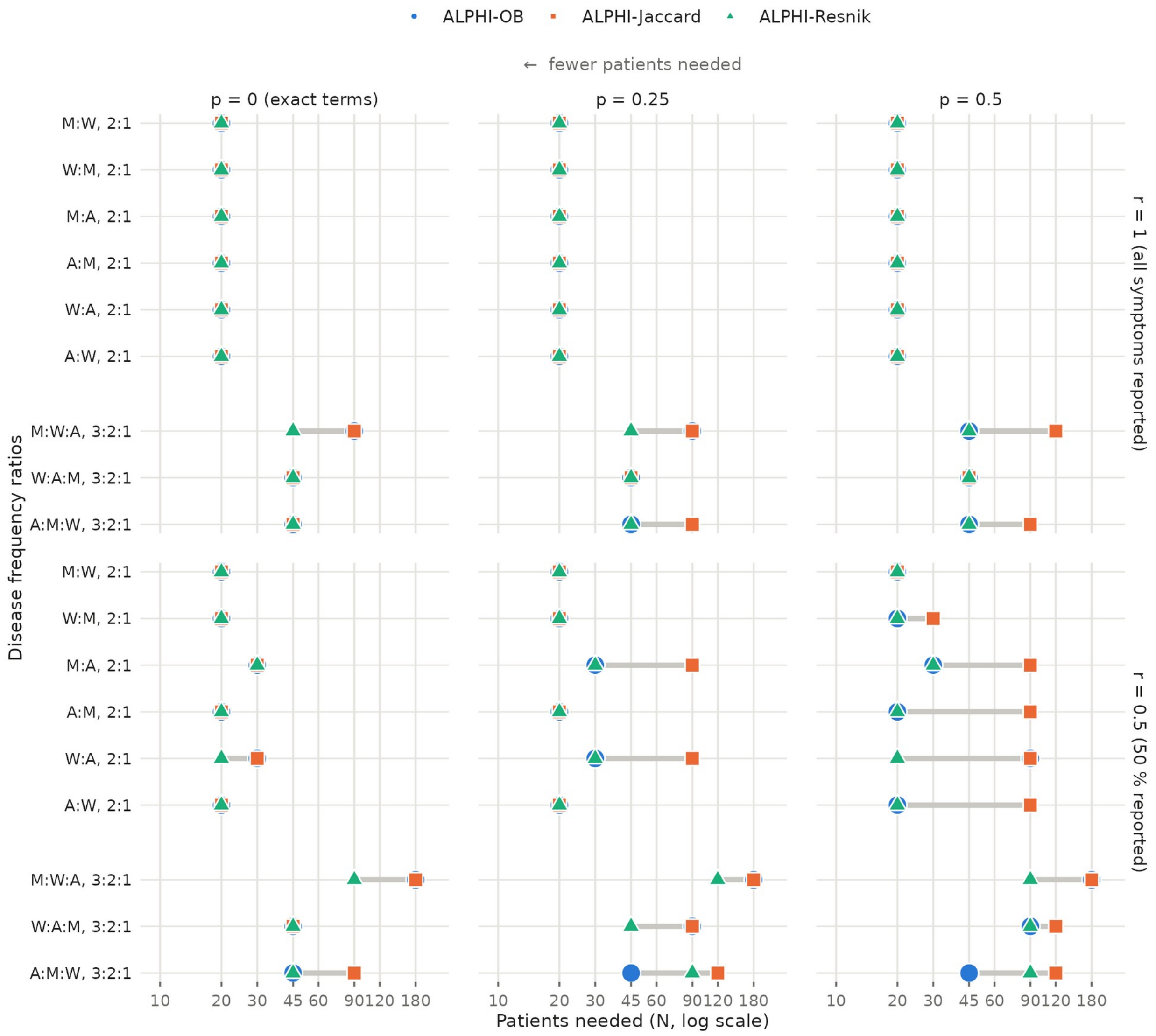


**Figure S3.** Number of patients required to separate distinct diseases in Simulation 3 by the three distances, without the latent class check. For each pair (2:1) or triple (3:2:1) of Marfan syndrome (M), Wilson disease (W) and Alagille syndrome (A), the smallest cohort size (log scale) at which the true number of diseases was found in at least 80% of runs, by reporting rate (r, rows: all or half of the symptoms reported) and term substitution probability (p, columns). The smallest size was found by bisection among cohort sizes of 10 to 180 patients. The values of the three distances (ALPHI-OB, ALPHI-Jaccard and ALPHI-Resnik) for one scenario are joined by a segment. Single-disease controls (N = 10–180) were divided in 6 of 730 runs with ALPHI-OB (5 flagged by the sparse-feature check), and in none of 720 with ALPHI-Jaccard or ALPHI-Resnik.

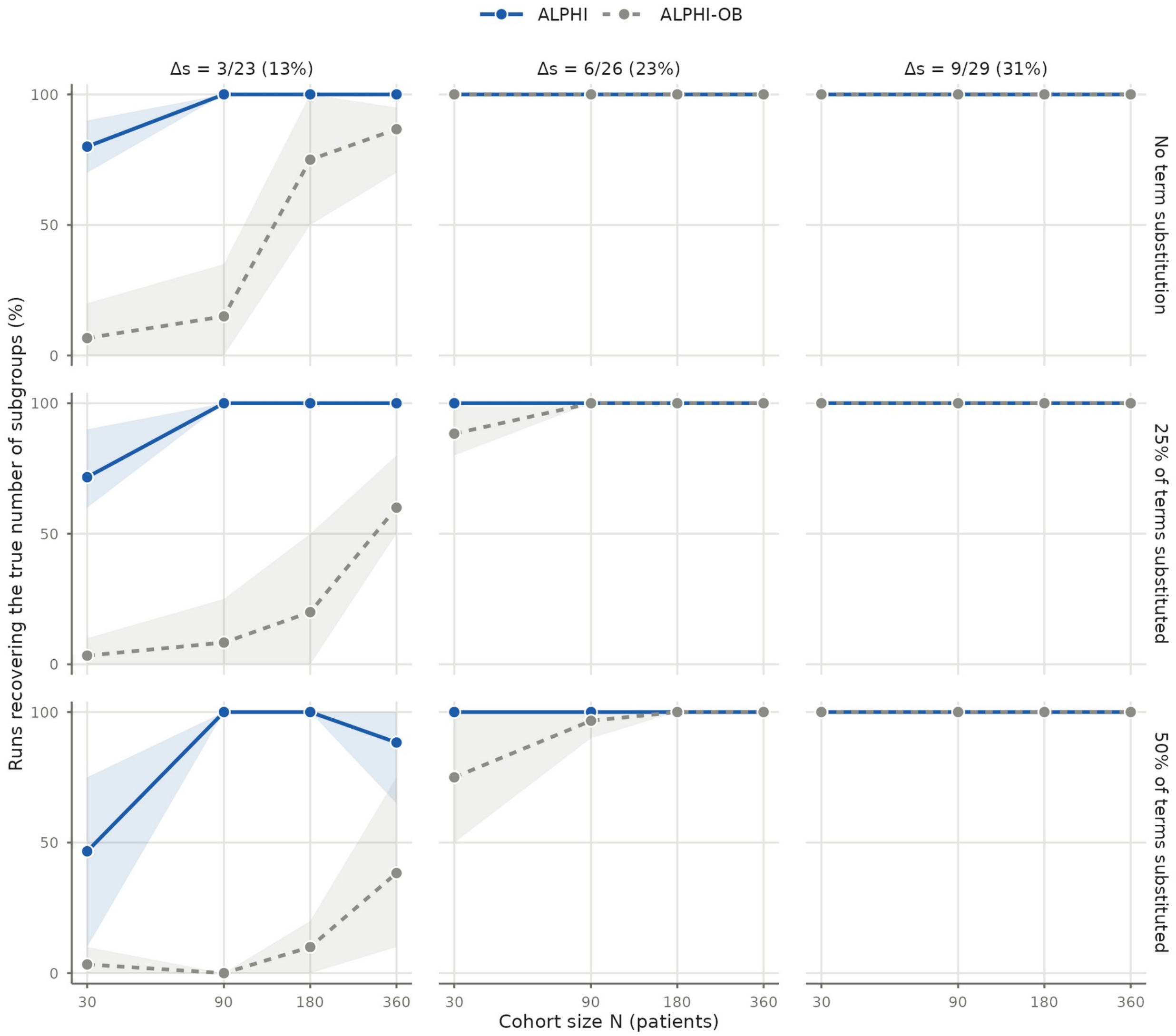


**Figure S4.** Effect of the latent class check on the recovery of two latent subgroups (D1S2) in Simulation 2. Percentage of cohorts in which the true number of subgroups was found by ALPHI (with the check; solid blue line) and ALPHI-OB (without the check; dashed grey line), by cohort size N, distinguishing symptoms (Δs, columns) and term substitution (none, 25% or 50% of terms substituted; rows). Δs: number of distinguishing symptoms of a subgroup over the number of symptoms defining it (20 core symptoms shared by all subgroups plus the distinguishing symptoms), with the percentage in parentheses. Lines: mean over compositions C1–C3; shaded bands: range over compositions. Dispersed and concentrated layouts are pooled; complete reporting. None of the 1,080 cohorts without subgroups (D1S0) was divided by ALPHI.

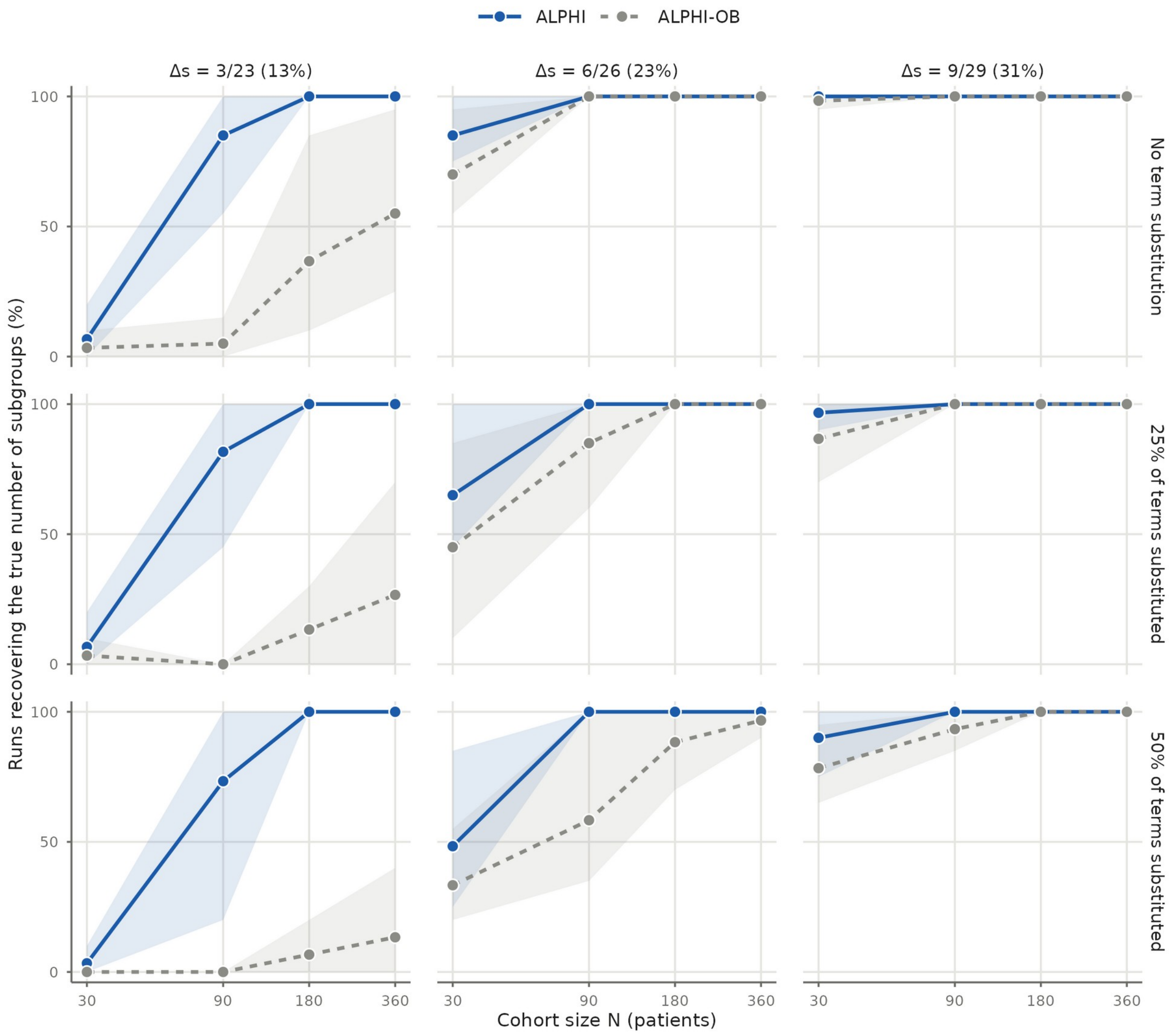


**Figure S5.** Effect of the latent class check on the recovery of three latent subgroups (D1S3) in Simulation 2. Percentage of cohorts in which the true number of subgroups was found by ALPHI (with the check; solid blue line) and ALPHI-OB (without the check; dashed grey line), by cohort size N, distinguishing symptoms (Δs, columns) and term substitution (none, 25% or 50% of terms substituted; rows). Δs: number of distinguishing symptoms of a subgroup over the number of symptoms defining it (20 core symptoms shared by all subgroups plus the distinguishing symptoms), with the percentage in parentheses. Lines: mean over compositions C1–C3; shaded bands: range over compositions. Dispersed and concentrated layouts are pooled; complete reporting. None of the 1,080 cohorts without subgroups (D1S0) was divided by ALPHI.

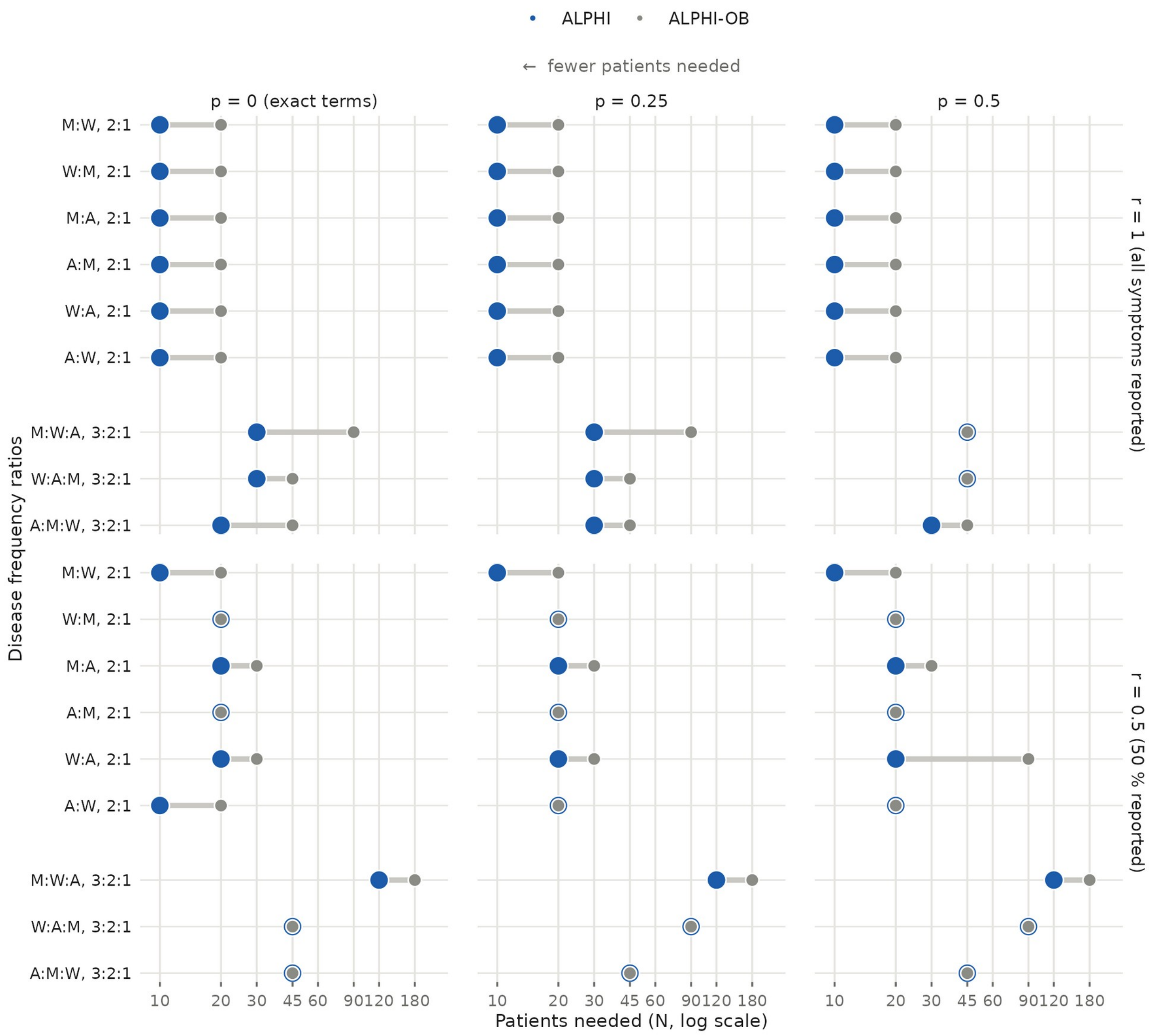


**Figure S6.** Effect of the latent class check on the number of patients required to separate distinct diseases in Simulation 3. For each pair (2:1) or triple (3:2:1) of Marfan syndrome (M), Wilson disease (W) and Alagille syndrome (A), the smallest cohort size (log scale) at which the true number of diseases was found in at least 80% of runs, by reporting rate (r, rows: all or half of the symptoms reported) and term substitution probability (p, columns). The smallest size was found by bisection among cohort sizes of 10 to 180 patients. Large blue dots: ALPHI (with the check); small grey dots: ALPHI-OB (without the check); the two values for one scenario are joined by a segment, and a grey dot inside a blue ring marks equal values. Single-disease controls were divided in 1 of 370 runs by ALPHI and 6 of 370 by ALPHI-OB with complete reporting, and in none of 360 by either with half of the symptoms reported.

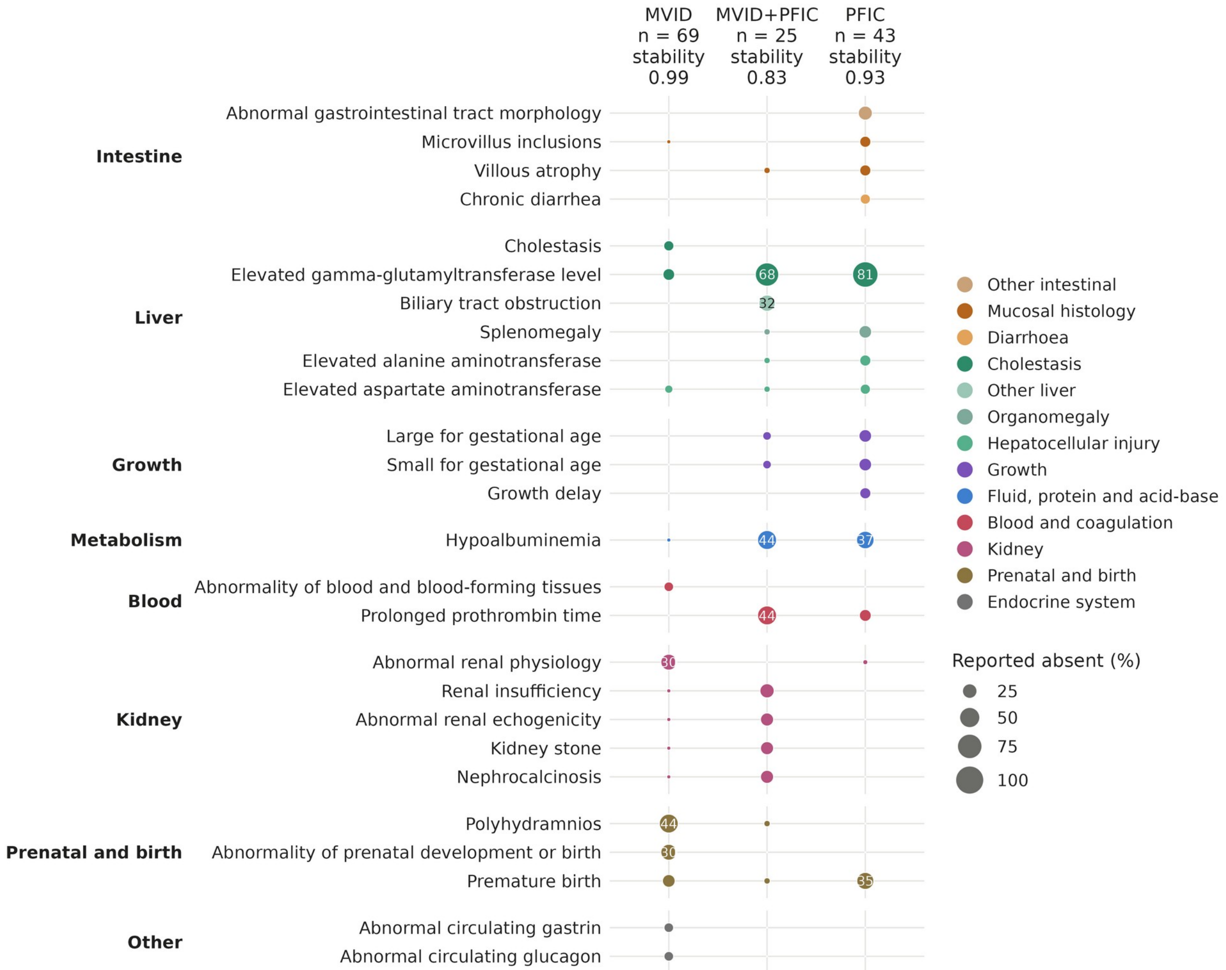


**Figure S7.** Symptoms reported absent in the three ALPHI clusters of the *MYO5B* cohort (MVID, 69 patients; MVID+PFIC, 25; PFIC, 43). Symptoms explicitly reported as absent were not used for clustering. A symptom was counted as absent when listed as absent and not as present (intermittent symptoms, recorded as both, were excluded). Terms reported absent in at least 10% of the patients of one cluster are shown, grouped by organ and colored by organ subgroup. Dot area and number: percentage of patients of the cluster. Stability: mean Jaccard index of each cluster over 50 subsamples of 80% of patients. HPO, Human Phenotype Ontology; MVID, microvillus inclusion disease; PFIC, progressive familial intrahepatic cholestasis.

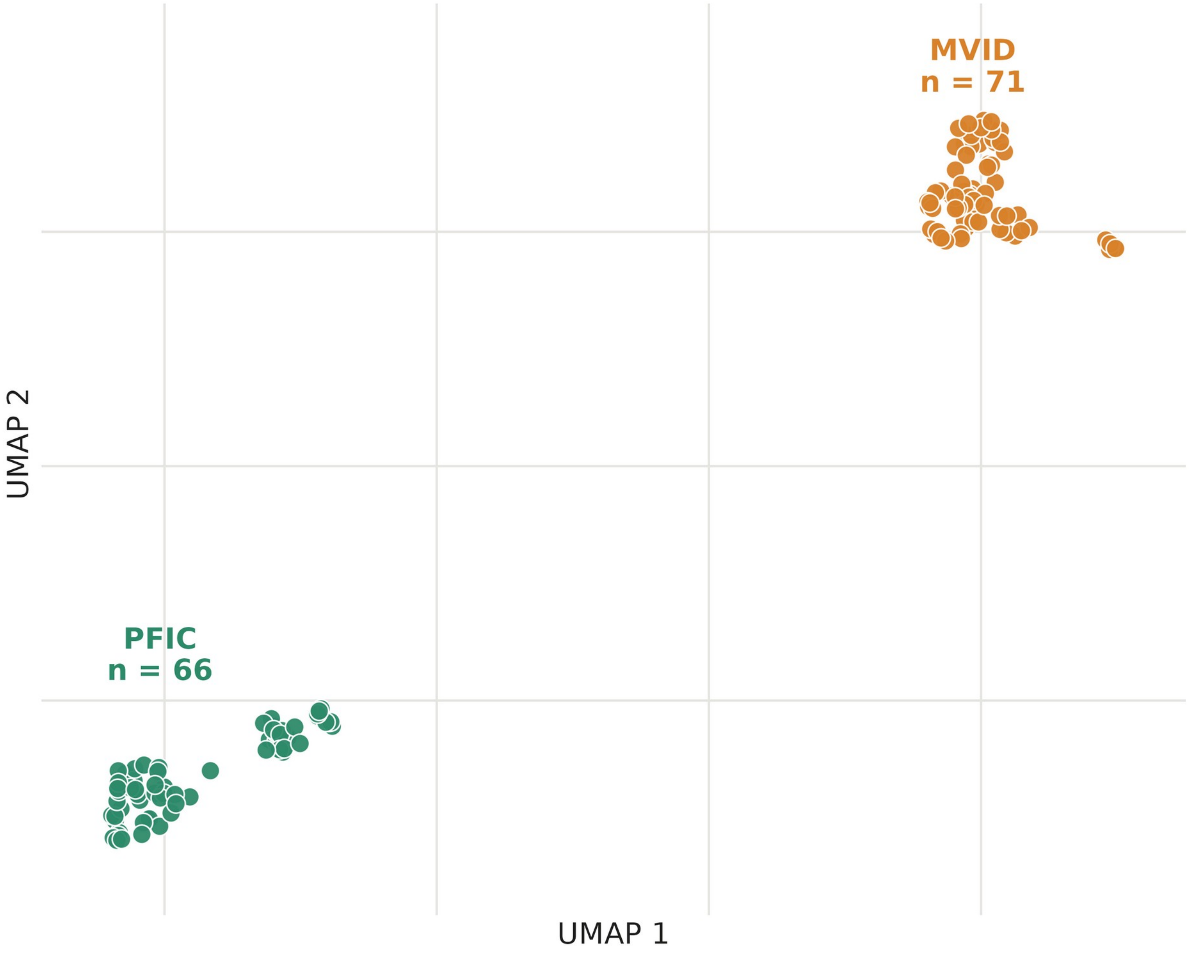


**Figure S8.** *MYO5B* patients in UMAP space, clustered with ALPHI-Resnik (without the latent class check). Two-dimensional uniform manifold approximation and projection (UMAP) of the Resnik distances between the 137 patients, colored by cluster: MVID (71 patients) and PFIC (66 patients). The two-dimensional embedding is for display; clusters were found in a ten-dimensional embedding of the same distances. MVID, microvillus inclusion disease; PFIC, progressive familial intrahepatic cholestasis.

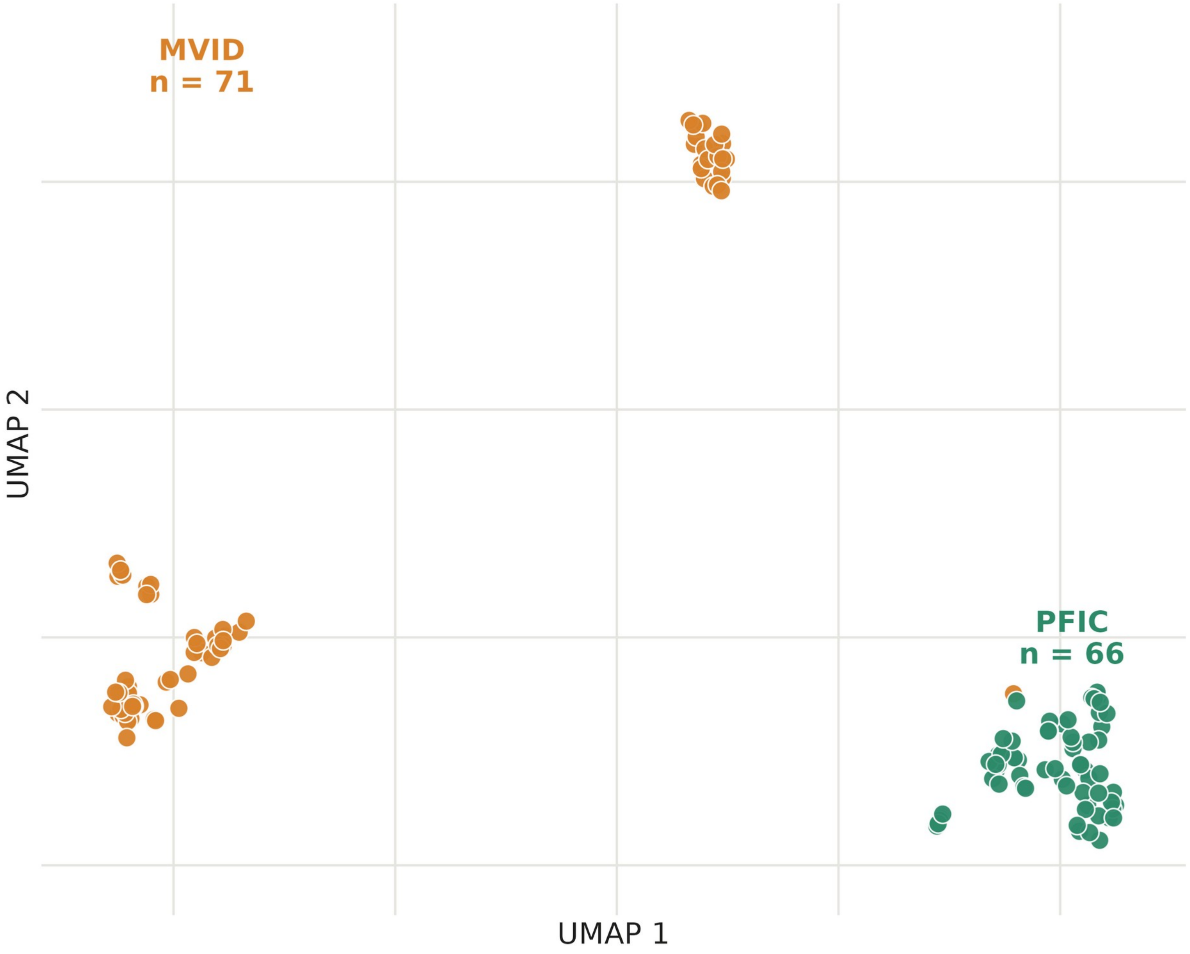


**Figure S9.** *MYO5B* patients in UMAP space, clustered with ALPHI-Jaccard (without the latent class check). Two-dimensional uniform manifold approximation and projection (UMAP) of the Jaccard distances between the 137 patients, colored by cluster: MVID (71 patients) and PFIC (66 patients). The two-dimensional embedding is for display; clusters were found in a ten-dimensional embedding of the same distances. MVID, microvillus inclusion disease; PFIC, progressive familial intrahepatic cholestasis.

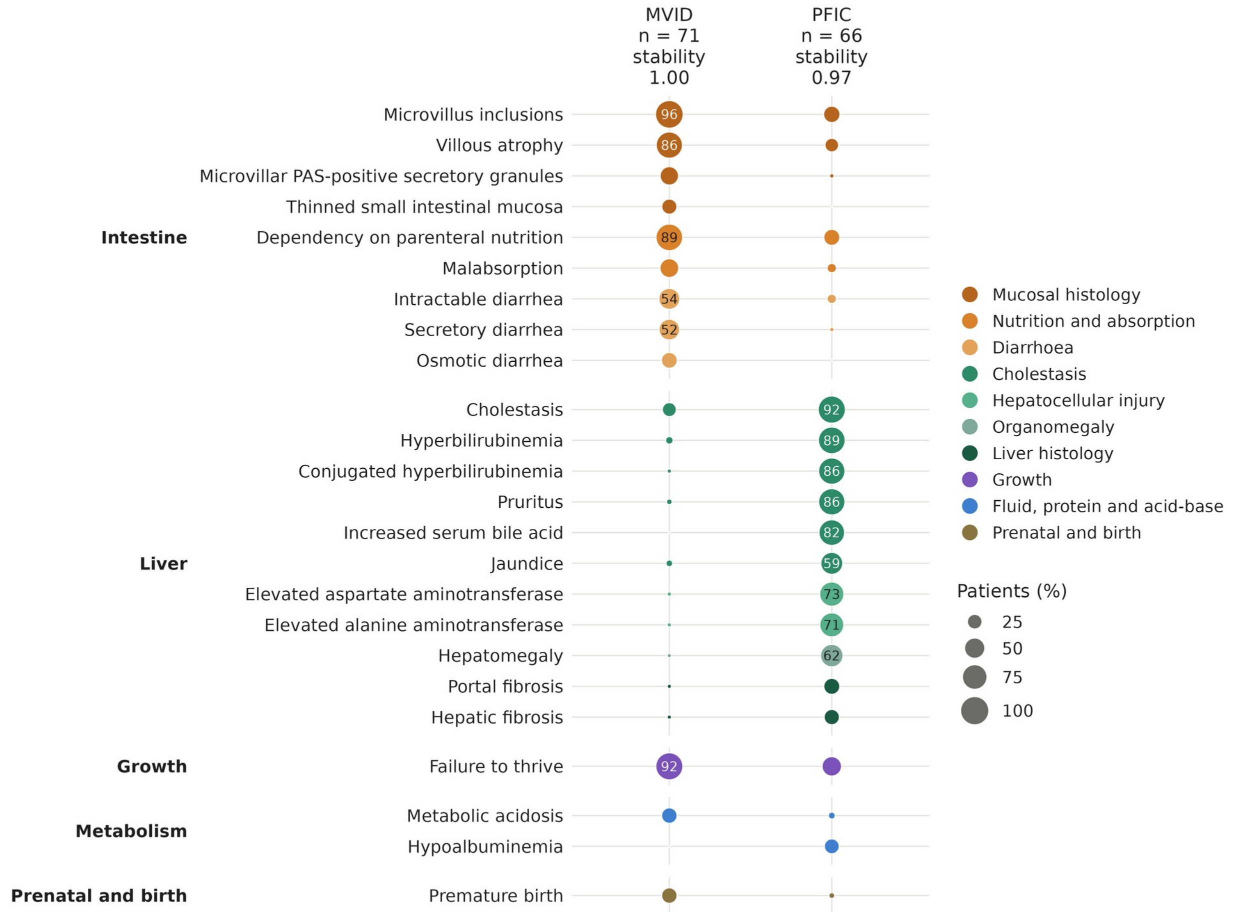


**Figure S10.** Phenotypic makeup of the *MYO5B* clusters obtained with ALPHI-Resnik (without the latent class check). Prevalence of the HPO terms whose frequency differs by at least 20 percentage points between the MVID (71 patients) and PFIC (66 patients) clusters, grouped by organ and colored by organ subgroup. Dot area and number: percentage of patients of the cluster. Stability: mean Jaccard index of each cluster over 50 subsamples of 80% of patients. HPO, Human Phenotype Ontology; MVID, microvillus inclusion disease; PFIC, progressive familial intrahepatic cholestasis.

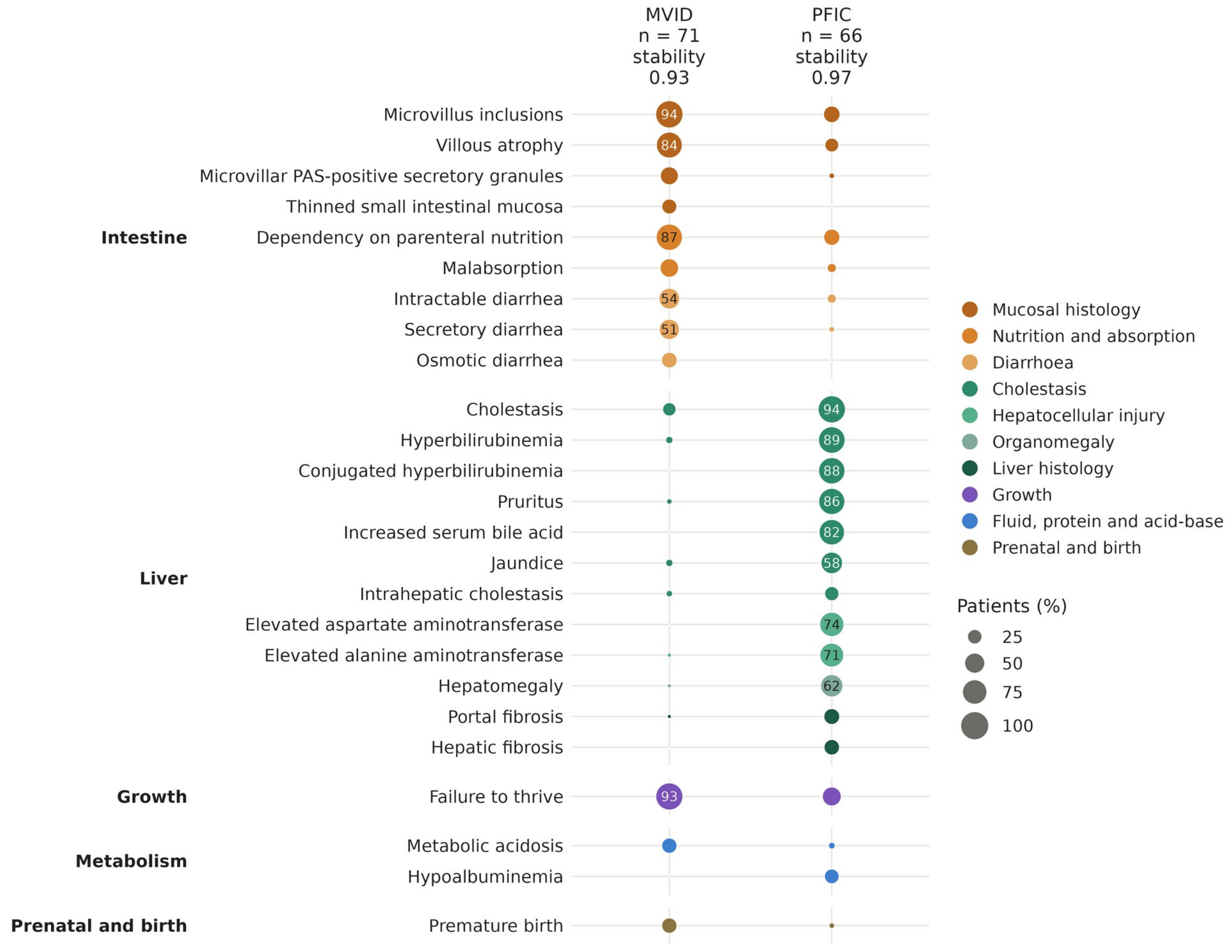


**Figure S11.** Phenotypic makeup of the *MYO5B* clusters obtained with ALPHI-Jaccard (without the latent class check). Prevalence of the HPO terms whose frequency differs by at least 20 percentage points between the MVID (71 patients) and PFIC (66 patients) clusters, grouped by organ and colored by organ subgroup. Dot area and number: percentage of patients of the cluster. Stability: mean Jaccard index of each cluster over 50 subsamples of 80% of patients. HPO, Human Phenotype Ontology; MVID, microvillus inclusion disease; PFIC, progressive familial intrahepatic cholestasis.

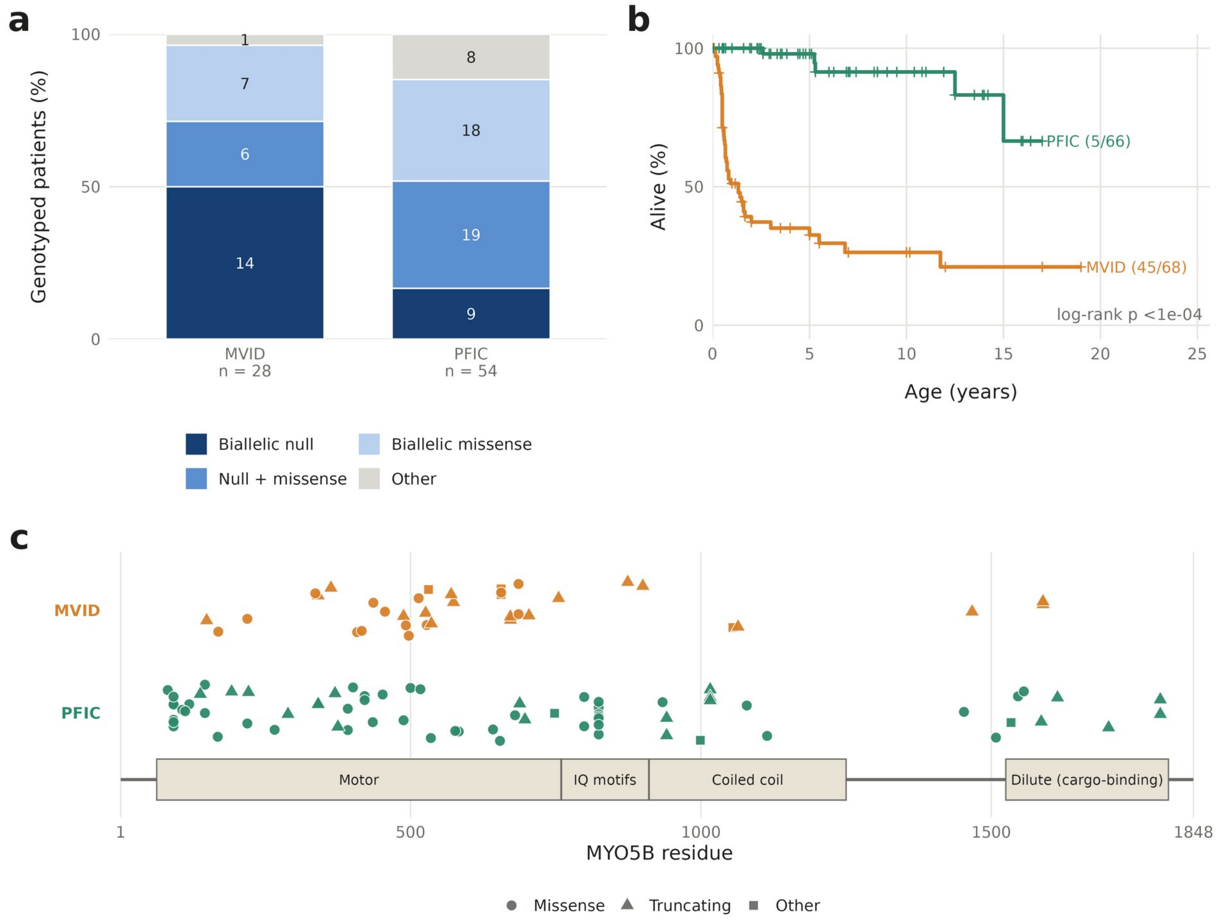


**Figure S12.** Genotype and survival of the *MYO5B* clusters obtained with ALPHI-Resnik (without the latent class check); neither was used for clustering. (A) Genotype class of patients with both variants classified (null: nonsense, frameshift or splice site). (B) Kaplan–Meier survival from birth; deaths/patients in parentheses; tick marks, censoring at last follow-up. (C) Position of variants on the MYO5B protein, one track per cluster (circles, missense; triangles, truncating; squares, other); splice variants without a protein position are not shown. Domain boundaries from UniProt Q9ULV0. MVID, microvillus inclusion disease; PFIC, progressive familial intrahepatic cholestasis.

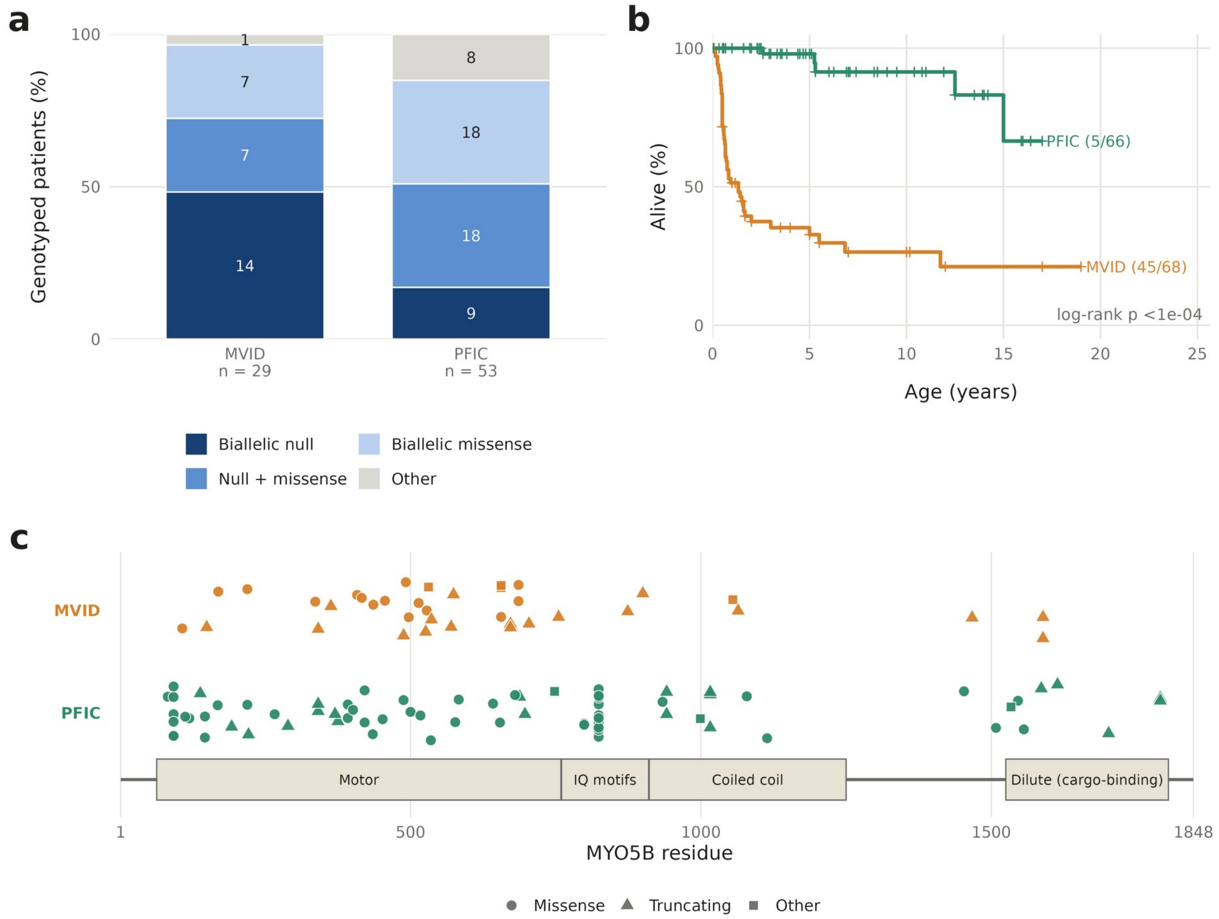


**Figure S13.** Genotype and survival of the *MYO5B* clusters obtained with ALPHI-Jaccard (without the latent class check); neither was used for clustering. (A) Genotype class of patients with both variants classified (null: nonsense, frameshift or splice site). (B) Kaplan–Meier survival from birth; deaths/patients in parentheses; tick marks, censoring at last follow-up. (C) Position of variants on the MYO5B protein, one track per cluster (circles, missense; triangles, truncating; squares, other); splice variants without a protein position are not shown. Domain boundaries from UniProt Q9ULV0. MVID, microvillus inclusion disease; PFIC, progressive familial intrahepatic cholestasis.
